# Dissection of the clinical phenome of major depressive disorder into subdomains and subgroups in relation to activated immune-inflammatory profiles

**DOI:** 10.1101/2025.10.21.25338439

**Authors:** Mengqi Niu, Yingqian Zhang, Xu Zhang, Chenkai Yangyang, Xiaoman Zhuang, Jing Li, Michael Maes

## Abstract

**Background:** Major depressive disorder (MDD) is a prevalent mood disorder characterized by low remission rates and a high risk of suicide. It encompasses various clinical domains, including depression, anxiety, vegetative, physiosomatic and melancholic symptoms, and suicidal behaviors.

**Aims:** To dissect the clinical phenome of MDD, identify potential constructs within the symptom domains, delineate MDD subclasses, investigate the predictive role of recurrence of illness (ROI) on the phenome, and analyze the relationship between those clinical features and immune activation biomarkers.

**Methods:** A total of 125 MDD patients and 40 healthy controls were recruited for this study. Assays included serum immunological features (M1, Th1, Th2, Th17, IRS, CIRS, and TNF signaling), negative acute phase proteins (APPs; transferrin, albumin), sCD40L, EGF, and Flt-3L.

**Results:** Bifactor analysis integrated multiple clinical domains into a general factor, labeled “OSOD” (overall severity of depression), while extracting a single group factor of physiosomatic symptoms. Cluster analysis identified two subgroups of MDD: major dysmood disorder (MDMD) and simple dysmood disorder (SDMD). ROI mediates the impact of adverse childhood experiences on OSOD and correlates with enhanced immune activation (increased Th17). Downregulation of negative APPs and Flt-3L alongside increased EGF and TNF signaling distinguish MDMD versus SDMD or MDD from controls with great accuracy. Up to 60% of MDD patients show immune aberrations with a specificity of 92.1%.

**Conclusion:** This study promotes a novel diagnostic approach that integrates ROI, OSOD, physiosomatic symptoms, MDMD versus SDMD, and immune profiling. It has the potential to establish the groundwork for future personalized therapeutic strategies.

## Introduction

Major Depressive Disorder (MDD) is a prevalent mood disorder characterized by high incidence, low remission rates, and elevated suicide risk (1). Globally, it stands as the second leading cause of disability, affecting over 3.32 million individuals (2). This disorder is characterized by a wide array of different symptom domains including affective (depression, anxiety), vegetative, physiosomatic (psychosomatic), chronic-fatigue-syndrome (CFS) and melancholia symptoms as well as suicidal behaviors (SB) including suicidal attempts (SA) and suicidal ideation (SI) (3–8). The aggregate of all those symptoms is labeled as the “phenome of MDD” (7).

A bifactorial method has recently been recommended for modeling clinical domains in depression (9). Certain studies indicated that bifactor models, incorporating a general factor alongside specific group factors for depression, anxiety, and somatic symptoms, may most accurately encapsulate the phenome of depression (10). A separate study indicated that a bifactor model, comprising a general factor and two subgroup factors—specifically a somatic factor and a cognitive component—yielded the optimal fit (11). Nonetheless, it remains unexamined which latent constructs may underlie the depression, anxiety, vegetative, psychosomatic, CFS-like, melancholic subdomains, and SB, or whether these dimensions should be considered distinct clinical constructs.

A large part (>50%) of patients with MDD show one or multiple recurrences of the depressive episodes and SB (12). This recurrence of illness (ROI) can be quantified by computing an ROI index based on self-reported or physician-rated depression episode frequency, alongside lifetime SI and SA (13). Adverse childhood experiences (ACEs) impact disease severity and disability, with ROI potentially serving as a mediator or indicator of this effect (13). Importantly, in Thai and Brazilian samples, ROI was shown to predict specific symptom presentations of the acute phase of severe MDD, including affective and physiosomatic symptoms, as well as overall depression severity (4–6, 14, 15). Moreover, studies conducted in Thailand and Brazil have confirmed a strong association between ACEs, ROI, these symptom dimensions, and current SB (7, 16–20). However, the relationships between ACEs, ROI, and these current phenome features remain unexplored in Chinese MDD patients.

A precision nomothetic psychiatry approach has further subtyped MDD into two distinct subclasses: major dysmood disorder (MDMD) and simple dysmood disorder (SDMD) (21). Compared to patients with SDMD, those with MDMD exhibit more severe symptoms of depression, anxiety, chronic fatigue, and SB, alongside a higher ROI index (21, 22). Whether the clinical characteristics (e.g., ROI and current phenome) of these two major MDD subtypes in China are similar to findings from other countries remains to be determined.

Different stages of MDD are associated with distinct alterations in the homeostatic balance between the immune-inflammatory response system (IRS) and the compensatory immune regulatory system (CIRS) (23). The IRS encompasses the macrophage M1 phenotype, T helper (Th)1, and Th17 profiles. IRS activation in MDD, especially MDMD, is characterized by elevated levels of pro-inflammatory cytokines, including IL-1, soluble IL-1 receptor antagonist (sIL-1RA), interferon (IFN)-γ, IL-2, IL-6, IL-12p70, and tumor necrosis factor (TNF)-α, as well as Th17 cell features including IL-17A (4, 24–26). This research has also identified elevated TNF signaling in MDMD, as evidenced by increased levels of TRAIL, TNF-α, and TNF-β. This pathway is actively involved in inflammation, cell death, and glial cell proliferation (22, 27). Concurrently, M1 activation during the acute phase of MDD can trigger an acute-phase response in the liver, manifested by decreased plasma concentrations of negative acute phase proteins (APPs) like albumin and transferrin (23). Some of these cytokines or activated T cells might induce neuroinflammation and exert central neurotoxic effects leading to neuronal dysfunctions or structural damage to central neurons (1).

Conversely, the CIRS controls excessive inflammation, promotes the restoration of homeostasis, and plays a healing role during recovery following IRS activation (23). CIRS activation is accompanied by the production of immunoregulatory cytokines from Th2 cells (IL-4 and IL-5) and T regulatory (Treg) cells (IL-10), which collectively attenuate M1 and Th1 polarization (23, 24, 26). Furthermore, certain immune receptor antagonists, such as sIL-1RA, indicate M1 activation while also exhibiting immunoregulatory effects (28).

Prior studies have revealed a significant relationship between ROI and those immune profiles, including M1, Th1, Th2, Th17, IRS, and CIRS (18). However, the precise relationship between these immune profiles and ROI and current phenome in Chinese MDD patients, as well as their context within different MDD subtypes (MDMD and SDMD), remains unclear, representing a significant knowledge gap.

Apart from the above classical immune profiles that play a role in the pathophysiology of the acute phase of MDMD (19, 22), there are other pro-inflammatory costimulatory molecules (e.g., soluble CD40 ligand or CD40L), immunoregulatory growth factors (e.g., epidermal growth factor or EGF), and immune system modulators (e.g., Fms-like tyrosine kinase 3 ligand or Flt-3L) that may be involved in MDD. CD40L is primarily expressed by activated CD4+ T cells and serves as an important costimulatory molecule in the immune system (29). Studies have found elevated numbers of CD40L+ T cells in MDD and increased expression of sCD40L is associated with its severity (25, 30). Controversy exists regarding peripheral EGF levels in depressed patients compared to healthy controls, with some studies reporting elevation and others reduction (31, 32). EGF is immunoregulatory by enhancing epithelial healing and regeneration, promoting tissue remodeling, barrier integrity and resolution of inflammation (33). Flt-3L is a crucial immune system development factor that primarily regulates the development of dendritic cells (DCs) by binding to its receptor, CD135 (34). Flt-3L enhances plasmacytoid DCs which produce Treg cells (producing tolerogenic cytokines) and play a role in conventional DCs promoting Treg induction and FoxP3 Treg differentiation and maintenance (35, 36). Nevertheless, the involvement of these three factors in MDD has remained elusive.

This study aims to dissect the phenome features of major depression to a) delineate which latent constructs underpin the symptom domains of MDD using bifactor analysis, b) examine whether MDD may be separated in the distinct subclasses, c) examine whether the clinical domains and subclasses are externally validated by the immune markers (IRS/CIRS profiles, negative APPs, sCD40L, EGF, and Flt-3L).

## Methods

### Participants

A total of 165 participants were recruited for this study, including 125 patients and 40 healthy controls (HCs). This study was a case-control cross-sectional design carried out at the Mental Health Center of Sichuan Provincial People’s Hospital in Chengdu, China. The participants’ ages varied from 18 to 65 years, exhibiting a balanced gender ratio. Patients with MDD were diagnosed based on DSM-5 criteria and exhibited a Hamilton Depression Rating Scale 21 (HAMD-21) score exceeding 18 (37). Additionally, patients were effectively categorized into two subgroups—MDMD and SDMD—through cluster analysis (see below). The control group was composed of hospital personnel, their family, and patients’ friends, matched to the case group according to age, gender, education, and body mass index (BMI). All participants or their legal guardians provided written informed consent. The Ethics Committee of Sichuan Provincial People’s Hospital approved the study [Ethics (Research) 2024-203].

Exclusion criteria for this study included: a) diagnosis of other major psychiatric disorders such as bipolar disorder, schizoaffective disorder, schizophrenia, psycho-organic disorders, substance use disorders (except nicotine dependence), and autism spectrum disorder; b) systemic conditions such as type 1 diabetes, rheumatoid arthritis, autoimmune disorders, systemic lupus erythematosus, inflammatory bowel disease, chronic obstructive pulmonary disease, psoriasis, or cancer; c) severe allergic reactions within the past month; d) being pregnant or breastfeeding; e) disorder related to development or personality, or neurological diseases such as Alzheimer’s disease, Parkinson’s disease, stroke, epilepsy, brain tumors, or multiple sclerosis; f) experiencing an infection during the last three months; g) surgery conducted within the past three months; h) present administration of immunosuppressants, corticosteroids, or other agents that modify the immune system; i) use of therapeutic amount of antioxidants or Omega-3 supplements in the past three months; j) frequent consumption of analgesics. Individuals in the control group were excluded if they had been diagnosed with MDD, dysthymia, DSM-5 anxiety disorders, or had a familial history of mood disorders, substance use disorders (except nicotine dependence), or suicide.

### Clinical Assessment

A qualified doctor performed a semi-structured interview to collect demographic information, medical history, mental history, and familial history. Validation of psychiatric diagnoses was conducted utilizing the Mini International Neuropsychiatric Interview (M.I.N.I.) (38), which evaluates various psychiatric conditions, including depressive episodes, hypomanic episodes, dysthymia, social anxiety disorder, panic disorder, generalized anxiety disorder, agoraphobia, post-traumatic stress disorder, obsessive-compulsive disorder, substance dependence/abuse (both alcohol and non-alcohol), psychotic disorders, eating disorders, and antisocial personality disorder.

On the same day, the assessor administered a variety of questionnaires to all participants to evaluate the severity of somatic-psychosomatic symptoms, anxiety, and depression. The severity of depression was evaluated using the HAMD-21 (37); Anxiety severity was measured with the Hamilton Anxiety Scale (HAMA) (39); Self-reported depression was quantified using the Beck Depression Inventory (40); and state anxiety was assessed through the State-Trait Anxiety Inventory (STAI) state subscale (41). The Somatic Symptom Scale-8 (SSS-8) evaluated the severity of somatic symptoms (e.g., pain, fatigue, digestive issues) that individuals had exhibited within the previous seven days. The FibroFatigue Scale (FFS) (42), a 12-item clinical interview instrument, was employed to measure the severity of symptoms associated with CFS (43). The Columbia Suicide Severity Rating Scale (C-SSRS) (44) evaluated both lifetime and current SA as well as SI. The clinical subdomains (e.g., vegetative, affective symptoms, physiosomatic symptoms, and overall severity of depression (OSOD) were developed using the above clinical scale scores; detailed computation methodologies are available in the **ESF, Table 1**.

**Table 1.**
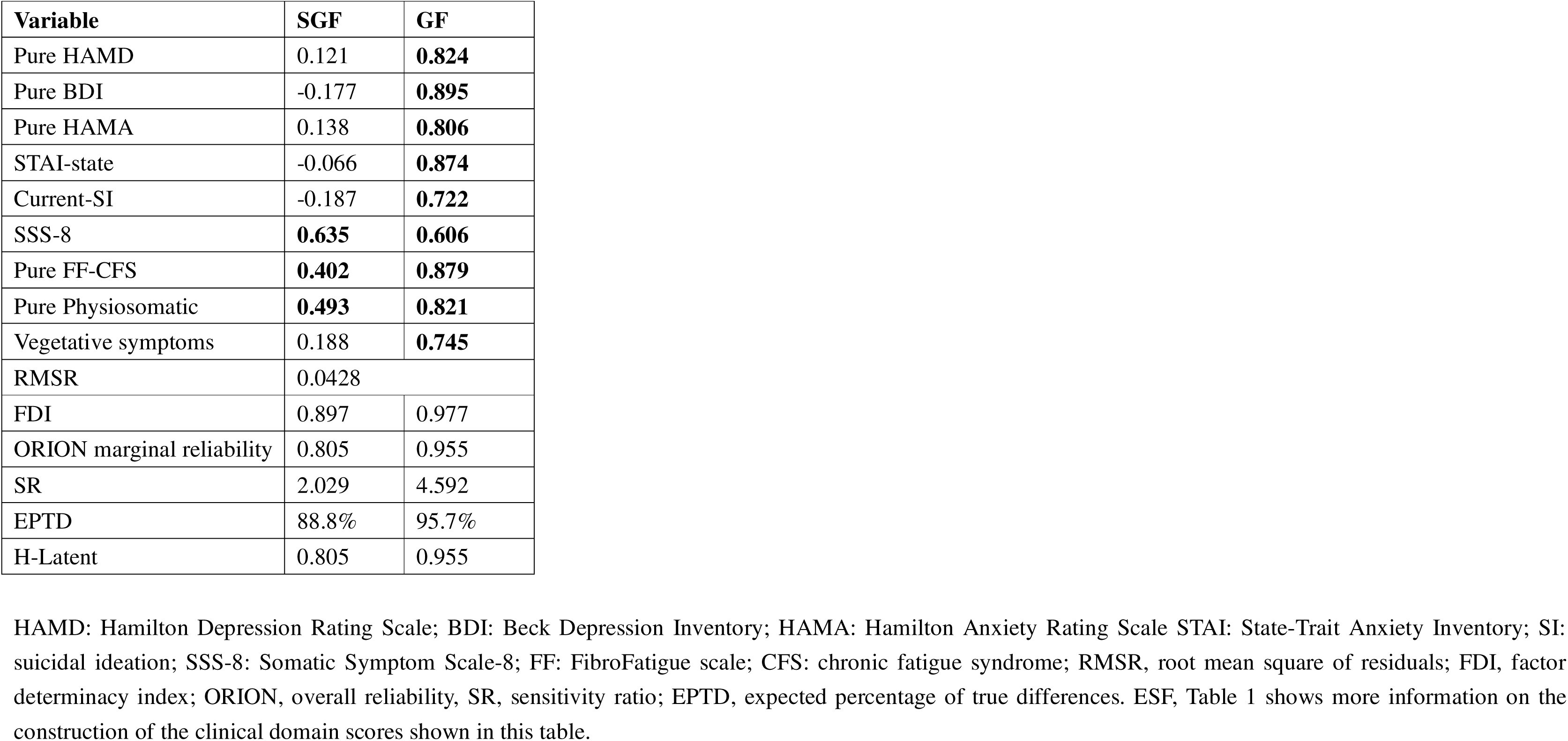
Exploratory bifactor loadings for the general factor (GF) and specific single group factor (SGF) of current phenome.

This study utilized two items from the C-SSRS to compute the recurrence of illness (ROI) index. The ROI is a composite score based on z-scores that is produced by adding the z-scores for the number of depressive episodes, SA, and SI across one’s lifetime (13). ACEs were evaluated utilizing the Childhood Trauma Questionnaire Short Form (CTQ-SF) (45), in a validated Chinese version (46). Scores were computed for five subscales: emotional abuse, physical abuse, sexual abuse, emotional neglect, and physical neglect (47).

This study examined the components of metabolic syndrome (MetS), which included height, weight, BMI, and waist circumference (WC). BMI was determined by dividing weight in kilograms by the square of height in meters. WC was assessed halfway between the iliac crest and the inferior rib. MetS was delineated in accordance with the 2009 joint statement from the American Heart Association and the National Heart, Lung, and Blood Institute (48). MetS was diagnosed upon the fulfillment of three or more of the following five criteria: (a) male WC ≥90 cm or female WC ≥80 cm; (b) triglycerides ≥150 mg/dL; (c) male HDL-C <40 mg/dL or female HDL-C <50 mg/dL; (d) systolic blood pressure ≥130 mm Hg or diastolic blood pressure ≥85 mm Hg, or administration of antihypertensive medications; (e) fasting glucose ≥100 mg/dL or confirmed diabetes mellitus. We utilized the MetS ranking in this study according to the number of MetS criteria employed.

### Assays

Blood samples were obtained between 06:30 and 08:00 hours, with 30 mL of fasting venous blood extracted using a reusable syringe and subsequently transferred into serum tubes. Following centrifugation at 3500 rpm, the serum was collected and aliquoted into Eppendorf tubes and stored at -80°C for later analysis. Serum cytokines were quantified utilizing Luminex xMAP technology on the Luminex 200 system (Luminex Corporation, Austin, TX, USA), employing the Human XL Cytokine Fixed Panel (catalog number: LKTM014B, Bio-Techne, R&D Systems), which measures cytokines, chemokines, and growth factors based on fluorescence intensity (FI) and concentration. Blank values were deducted to adjust for FI during measurements. The procedures involved: 1) dilute the sample using Calibrator Diluent RD6-65 (2x); 2) add 50 μL of the diluted sample and 50 μL of the bead mixture to each well, and incubate at room temperature with shaking at 850 rpm for 2 hours; 3) wash thrice, add 50 μL of diluted biotin-antibody combination, and incubate with shaking at 850 rpm for 1 hour; 4) wash again, add 50 μL of diluted streptavidin-PE, and incubate with shaking at 850 rpm for 30 minutes; 5) perform a final wash, add 100 μL of wash buffer to resuspend beads, and shake for 2 minutes. The Luminex 200 system was subsequently employed to quantify the 46 analytes. The intra-assay coefficient of variation for all analytes was less than 5%, while the inter-assay coefficient of variation was below 11.2%.

Serum transferrin was quantified with an immunoturbidimetric assay (DIAYS) on the ADVIA 2400, exhibiting a sensitivity of 0.03 g/L and intra-/inter-assay coefficients of variation of 1.96% and 0.67%, respectively. Serum albumin was quantified with the bromocresol green technique on the ADVIA 2400, exhibiting intra-/inter-assay coefficients of variation of 1.20% and 2.10%, respectively.

Comprehensive immune system protein data, encompassing cytokines, negative APPs (transferrin, albumin), sCD40L, EGF, and Flt-3L can be found in **ESF, Table 2**. Details about several immunological profiles (M1, Th1, Th2, Th17, IRS, CIRS, and TNF signaling) are available in the **ESF, Table 3**.

**Table 2.**
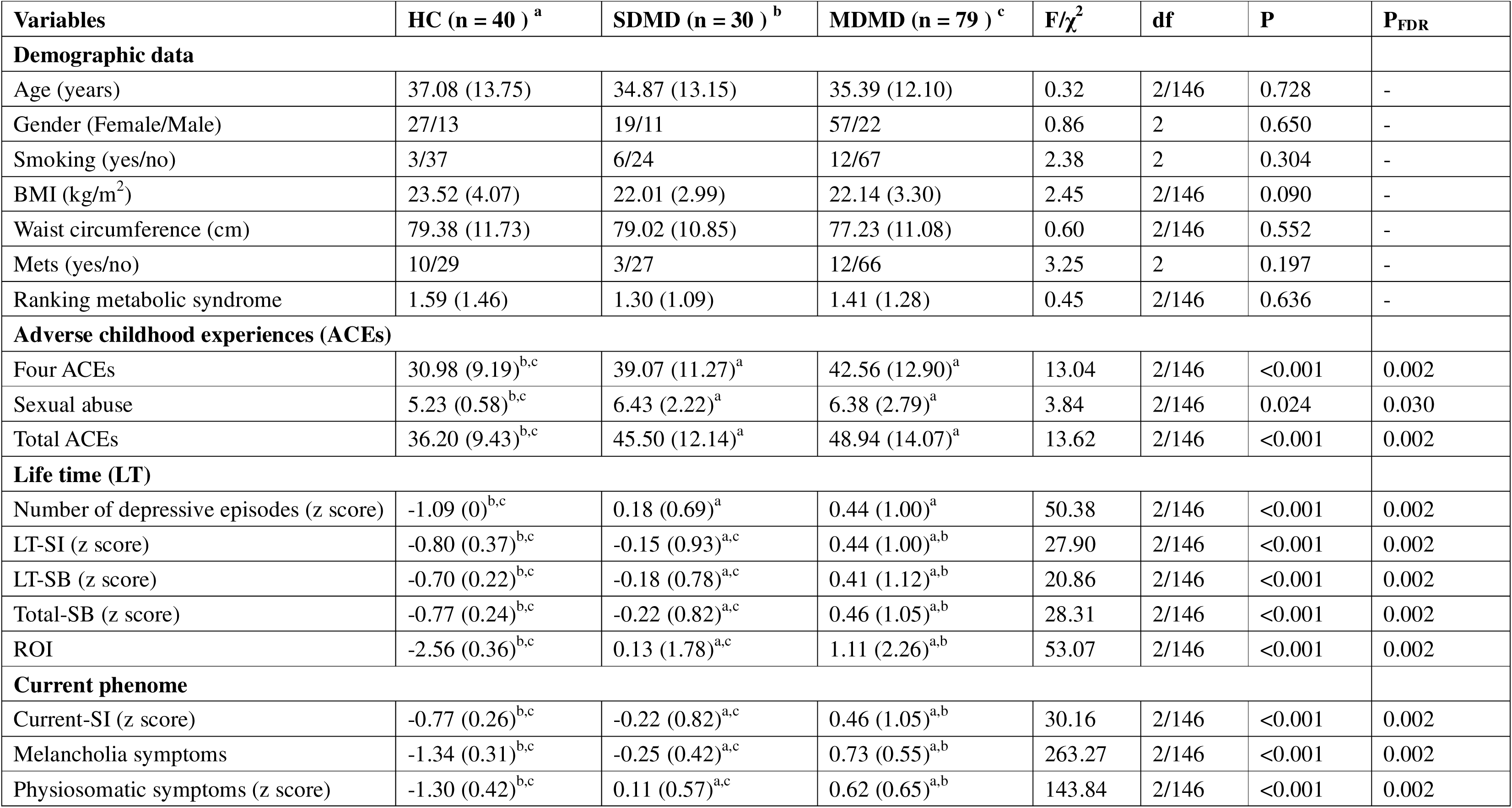
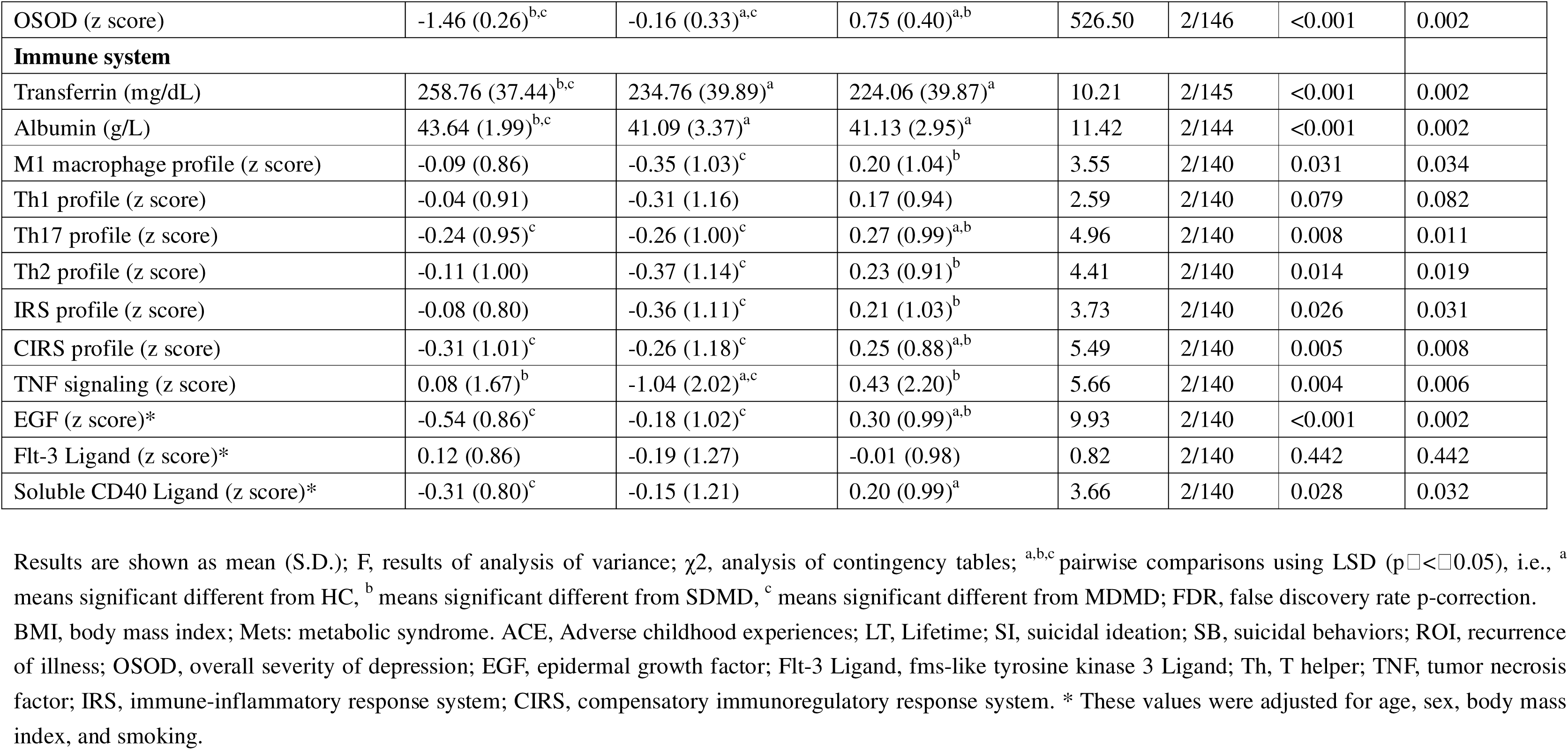
Features of patients with major dysmood disorder (MDMD) versus simple dysmood disorder (SDMD) and healthy controls (HC).

| Variables | HC (n = 40 ) <sup>a</sup> | SDMD (n = 30 ) <sup>b</sup> | MDMD (n = 79 ) <sup>c</sup> | F/$\chi^2$ | df | P | P<sub>FDR</sub> |
| --- | --- | --- | --- | --- | --- | --- | --- |
| **Demographic data** |  |  |  |  |  |  |  |
| Age (years) | 37.08 (13.75) | 34.87 (13.15) | 35.39 (12.10) | 0.32 | 2/146 | 0.728 | - |
| Gender (Female/Male) | 27/13 | 19/11 | 57/22 | 0.86 | 2 | 0.650 | - |
| Smoking (yes/no) | 3/37 | 6/24 | 12/67 | 2.38 | 2 | 0.304 | - |
| BMI (kg/m<sup>2</sup>) | 23.52 (4.07) | 22.01 (2.99) | 22.14 (3.30) | 2.45 | 2/146 | 0.090 | - |
| Waist circumference (cm) | 79.38 (11.73) | 79.02 (10.85) | 77.23 (11.08) | 0.60 | 2/146 | 0.552 | - |
| Mets (yes/no) | 10/29 | 3/27 | 12/66 | 3.25 | 2 | 0.197 | - |
| Ranking metabolic syndrome | 1.59 (1.46) | 1.30 (1.09) | 1.41 (1.28) | 0.45 | 2/146 | 0.636 | - |
| **Adverse childhood experiences (ACEs)** |  |  |  |  |  |  |  |
| Four ACEs | 30.98 (9.19)<sup>b,c</sup> | 39.07 (11.27)<sup>a</sup> | 42.56 (12.90)<sup>a</sup> | 13.04 | 2/146 | <0.001 | 0.002 |
| Sexual abuse | 5.23 (0.58)<sup>b,c</sup> | 6.43 (2.22)<sup>a</sup> | 6.38 (2.79)<sup>a</sup> | 3.84 | 2/146 | 0.024 | 0.030 |
| Total ACEs | 36.20 (9.43)<sup>b,c</sup> | 45.50 (12.14)<sup>a</sup> | 48.94 (14.07)<sup>a</sup> | 13.62 | 2/146 | <0.001 | 0.002 |
| **Life time (LT)** |  |  |  |  |  |  |  |
| Number of depressive episodes (z score) | -1.09 (0)<sup>b,c</sup> | 0.18 (0.69)<sup>a</sup> | 0.44 (1.00)<sup>a</sup> | 50.38 | 2/146 | <0.001 | 0.002 |
| LT-SI (z score) | -0.80 (0.37)<sup>b,c</sup> | -0.15 (0.93)<sup>a,c</sup> | 0.44 (1.00)<sup>a,b</sup> | 27.90 | 2/146 | <0.001 | 0.002 |
| LT-SB (z score) | -0.70 (0.22)<sup>b,c</sup> | -0.18 (0.78)<sup>a,c</sup> | 0.41 (1.12)<sup>a,b</sup> | 20.86 | 2/146 | <0.001 | 0.002 |
| Total-SB (z score) | -0.77 (0.24)<sup>b,c</sup> | -0.22 (0.82)<sup>a,c</sup> | 0.46 (1.05)<sup>a,b</sup> | 28.31 | 2/146 | <0.001 | 0.002 |
| ROI | -2.56 (0.36)<sup>b,c</sup> | 0.13 (1.78)<sup>a,c</sup> | 1.11 (2.26)<sup>a,b</sup> | 53.07 | 2/146 | <0.001 | 0.002 |
| **Current phenome** |  |  |  |  |  |  |  |
| Current-SI (z score) | -0.77 (0.26)<sup>b,c</sup> | -0.22 (0.82)<sup>a,c</sup> | 0.46 (1.05)<sup>a,b</sup> | 30.16 | 2/146 | <0.001 | 0.002 |
| Melancholia symptoms | -1.34 (0.31)<sup>b,c</sup> | -0.25 (0.42)<sup>a,c</sup> | 0.73 (0.55)<sup>a,b</sup> | 263.27 | 2/146 | <0.001 | 0.002 |
| Physiosomatic symptoms (z score) | -1.30 (0.42)<sup>b,c</sup> | 0.11 (0.57)<sup>a,c</sup> | 0.62 (0.65)<sup>a,b</sup> | 143.84 | 2/146 | <0.001 | 0.002 |
| OSOD (z score) | -1.46 (0.26)<sup>b,c</sup> | -0.16 (0.33)<sup>a,c</sup> | 0.75 (0.40)<sup>a,b</sup> | 526.50 | 2/146 | <0.001 | 0.002 |
| **Immune system** |  |  |  |  |  |  |  |
| Transferrin (mg/dL) | 258.76 (37.44)<sup>b,c</sup> | 234.76 (39.89)<sup>a</sup> | 224.06 (39.87)<sup>a</sup> | 10.21 | 2/145 | <0.001 | 0.002 |
| Albumin (g/L) | 43.64 (1.99)<sup>b,c</sup> | 41.09 (3.37)<sup>a</sup> | 41.13 (2.95)<sup>a</sup> | 11.42 | 2/144 | <0.001 | 0.002 |
| M1 macrophage profile (z score) | -0.09 (0.86) | -0.35 (1.03)<sup>c</sup> | 0.20 (1.04)<sup>b</sup> | 3.55 | 2/140 | 0.031 | 0.034 |
| Th1 profile (z score) | -0.04 (0.91) | -0.31 (1.16) | 0.17 (0.94) | 2.59 | 2/140 | 0.079 | 0.082 |
| Th17 profile (z score) | -0.24 (0.95)<sup>c</sup> | -0.26 (1.00)<sup>c</sup> | 0.27 (0.99)<sup>a,b</sup> | 4.96 | 2/140 | 0.008 | 0.011 |
| Th2 profile (z score) | -0.11 (1.00) | -0.37 (1.14)<sup>c</sup> | 0.23 (0.91)<sup>b</sup> | 4.41 | 2/140 | 0.014 | 0.019 |
| IRS profile (z score) | -0.08 (0.80) | -0.36 (1.11)<sup>c</sup> | 0.21 (1.03)<sup>b</sup> | 3.73 | 2/140 | 0.026 | 0.031 |
| CIRS profile (z score) | -0.31 (1.01)<sup>c</sup> | -0.26 (1.18)<sup>c</sup> | 0.25 (0.88)<sup>a,b</sup> | 5.49 | 2/140 | 0.005 | 0.008 |
| TNF signaling (z score) | 0.08 (1.67)<sup>b</sup> | -1.04 (2.02)<sup>a,c</sup> | 0.43 (2.20)<sup>b</sup> | 5.66 | 2/140 | 0.004 | 0.006 |
| EGF (z score)* | -0.54 (0.86)<sup>c</sup> | -0.18 (1.02)<sup>c</sup> | 0.30 (0.99)<sup>a,b</sup> | 9.93 | 2/140 | <0.001 | 0.002 |
| Flt-3 Ligand (z score)* | 0.12 (0.86) | -0.19 (1.27) | -0.01 (0.98) | 0.82 | 2/140 | 0.442 | 0.442 |
| Soluble CD40 Ligand (z score)* | -0.31 (0.80)<sup>c</sup> | -0.15 (1.21) | 0.20 (0.99)<sup>a</sup> | 3.66 | 2/140 | 0.028 | 0.032 |

Results are shown as mean (S.D.); F, results of analysis of variance; $\chi^2$ , analysis of contingency tables; <sup>a,b,c</sup> pairwise comparisons using LSD ( $p \leq 0.05$ ), i.e., <sup>a</sup> means significant different from HC, <sup>b</sup> means significant different from SDMD, <sup>c</sup> means significant different from MDMD; FDR, false discovery rate p-correction. BMI, body mass index; Mets: metabolic syndrome. ACE, Adverse childhood experiences; LT, Lifetime; SI, suicidal ideation; SB, suicidal behaviors; ROI, recurrence of illness; OSOD, overall severity of depression; EGF, epidermal growth factor; Flt-3 Ligand, fms-like tyrosine kinase 3 Ligand; Th, T helper; TNF, tumor necrosis factor; IRS, immune-inflammatory response system; CIRS, compensatory immunoregulatory response system. \* These values were adjusted for age, sex, body mass index, and smoking.

**Table 3.** Correlation matrix between adverse childhood experiences (ACEs), number of depressive episodes, the recurrence of illness (ROI) index and clinical domains of depression.

| Total (n = 165) | Variables | Total ACEs | Number of depressive episodes | ROI |
| --- | --- | --- | --- | --- |
|  | ROI | 0.547 (<0.001) ** | 0.751 (<0.001) ** | - |
|  | Total-SB | 0.537 (<0.001) ** | 0.513 (<0.001) ** | 0.873 (<0.001) ** |
|  | Current-SI | 0.483 (<0.001) ** | 0.472 (<0.001) ** | 0.743 (<0.001) ** |
|  | Melancholia symptoms | 0.449 (<0.001) ** | 0.565 (<0.001) ** | 0.637 (<0.001) ** |
|  | Physiosomatic symptoms | 0.441 (<0.001) ** | 0.615 (<0.001) ** | 0.635 (<0.001) ** |
|  | OSOD | 0.476 (<0.001) ** | 0.677 (<0.001) ** | 0.717 (<0.001) ** |
|  | Variables | Total ACEs | Number of depressive episodes | ROI |
| MDD (n = 125 ) | ROI | 0.462 (<0.001) ** | 0.595 (<0.001) ** | - |
|  | Total-SB | 0.471 (<0.001) ** | 0.330 (<0.001) ** | 0.847 (<0.001) ** |
|  | Current-SI | 0.396 (<0.001) ** | 0.271 (0.003) ** | 0.659 (<0.001) ** |
|  | Melancholia symptoms | 0.248 (0.007) ** | 0.149 (0.113) | 0.298 (0.001) ** |
|  | Physiosomatic symptoms | 0.249 (0.009) ** | 0.262 (0.006) ** | 0.287 (0.003) ** |
|  | OSOD | 0.319 (<0.001) ** | 0.341 (<0.001) ** | 0.448 (<0.001) ** |

ACE, Adverse childhood experiences; SI, suicidal ideation; SB, suicidal behaviors; ROI, recurrence of illness; OSOD, overall severity of depression. \*, correlation is significant at the 0.05 level; \*\*, correlation is significant at the 0.01 level.

### Data Analysis

Exploratory factor analysis was conducted utilizing Factor software (version 12.06.08 for Windows), including 1-factor, 2-factor, and 3-factor solutions, in addition to Schmid-Leiman and bifactor models. The selected final model is based on RMSR, factor quality, parallel analysis, and dimensional evaluation. In addition, a bifactor model should demonstrate that the variables exhibit significant loadings on the general factor (GF) and on a single group factor (SGF).

All remaining statistical analyses were conducted utilizing IBM SPSS for Windows (version 30). Two-step cluster analysis was performed to determine MDMD and SDMD subclasses based on current phenome severity scores (without ROI, suicidal behaviors, and immune biomarkers). Our method utilized the Schwarz Bayesian clustering criterion and the log-likelihood distance measure. The generated clusters were considered satisfactory when the silhouette measure of cohesiveness and separation attained a minimum of 0.5. Principal component analysis (PCA) was employed to derive principal components (PCs) reflecting specific symptom domains from the clinical dataset. The PC model was approved only when the Kaiser-Meyer-Olkin (KMO) measure of sample adequacy exceeded 0.7, Bartlett’s test of sphericity was significant, all loadings of the variables were > 0.7, the unidimensional criterion Unidimensional Congruence (UniCo) surpassed 0.95, and the explained common variance (ECV) criterion surpassed 0.85 (both UniCo and ECV indicating a unidimensional PC). Consequently, PC scores were calculated and utilized in further statistical studies.

Comparisons of continuous variables across groups were conducted using analysis of variance (ANOVA), and the relationship between categorical variables was evaluated by contingency table analysis. Pearson correlations assessed relationships among continuous variables, while point-biserial correlations evaluated links between continuous and binary variables. The false discovery rate (FDR) was used to adjust for multiple comparisons. Manual multiple regression analysis was employed to assess the influence of explanatory variables (e.g., demographic characteristics, ACEs, cytokine profiles, negative acute-phase reactants, and solitary immune modulatory cytokines/growth factors) on dependent variables (e.g., episodes, ROI, SB, affective symptoms, psychosomatic symptoms, OSOD, etc.). In addition, an automatic forward stepwise regression technique was utilized with a p-value criterion of 0.05 for the inclusion of variables. The automated modeling procedure employed entry and removal thresholds of p = 0.05 and p = 0.07, respectively. The regression outcomes encompassed standardized β coefficients, degrees of freedom, p-values, R^2^, and F-statistics. Heteroscedasticity was assessed utilizing White’s test and the modified Breusch-Pagan test. Multicollinearity was assessed by tolerance and variance inflation factors. All analyses were conducted as two-tailed tests with a significance level (α) established at 0.05. A binary logistic regression model was employed to compare MDD with the control group and MDMD with the SDMD group. The dependent variable was diagnosis (MDD or MDMD), with the control group or SDMD serving as the reference variable. Covariates comprised gender, WC, BMI, and education years. The results comprised Nagelkerke pseudo R^2^, p-values for the Wald statistic, odds ratios (OR), 95% confidence intervals (CI), regression coefficients (B), and standard errors (SE). Over-sampling (random replication) was implemented for the SDMD vs. MDMD and control vs. MDD groups to rectify subgroup imbalances, including the smaller sample sizes in the SDMD and control groups. The classification model’s accuracy was assessed by linear discriminant analysis combined with 10-fold cross-validation. The model’s performance was encapsulated by the area under the curve (AUC) of the receiver operating characteristic (ROC) curve, Gini index, maximum Kolmogorov-Smirnov (Max K-S) value, and overall model quality. Data transformations, such as log10, square root, rank, or Winsorization, were utilized, as necessary.

## Results

### Results of exploratory bifactorial analysis

To delineate the symptom dimensions underpinning the symptom domains of MDD, we performed different types of exploratory factor analysis including factor solutions with 1, 2 or 3 factors, and Schmid-Leiman and bifactor models. As shown in **Table 1**, 9 symptom domains were entered in the factor analysis. Melancholia was excluded from this analysis due to its substantial overlap with other symptom domains. The best model (in terms of RMSR, factor quality data, parallel analysis to determine the number of factors, and dimensionality assessment) is shown in **Table 1**. The results show that a bifactor model best fitted the data. All 9 variables entered in the analysis showed high loadings on the general factor (GF), indicating that the GF effectively explained most of the common variance. Nevertheless, a single group factor (SGF) consisting of SSS8, pure FF-CFS, and pure physiosomatic symptoms was retrieved and showed adequate model quality data. The overall model fit was adequate (RMSR = 0.0428, below Kelley’s criterion of 0.0778). **Table 1** shows that the Factor Determinacy Indices (FDI) and ORION marginal reliability metrics were adequate for both factors, indicating that the latent factor estimates of GF and SGF showed good determinacy and reliability. In addition, the Sensitivity Ratio (SR) and the Expected Percentage of True Differences (EPTD) further supported the validity of the model. Finally, both Generalized H index (H-Latent) values exceeded the 0.80 threshold, suggesting that GF and SGF are well-defined latent variables likely to remain stable across studies. Consequently, we have computed two different dimensional symptom scores which reflect this bifactorial model: a) the overall severity of depression (labelled OSOD) score (see **Table 2**) which reflects the severity of the GF and is computed as a PC extracted from all symptom domains, except current SI, which was entered separately in the analysis; and b) an index reflecting the SGF, computed as a PC extracted from the three somatic symptom domains that determine the SGF (labeled: Physiosomatic dimension).

### Results of cluster analysis and features of SDMD, MDMD and HC

To detect relevant subclasses in the data set, we performed a two-step cluster analysis. Based on MDD and the symptom domain scores of the current phenome, including melancholia, physiosomatic and OSOD scores (lifetime clinical data, ROI, ACEs and biomarkers were not included), cluster analysis generated three classes with great accuracy, as shown by an overall silhouette index of 0.70. The first cluster-generated class (n = 40) corresponded to the normal controls, whereas the MDD sample was divided into two subsamples, a first cluster comprising 30 patients (cluster 1) and a second cluster comprising 79 subjects (cluster 2). The analysis was unable to classify 16 subjects.

**Table 2** shows the features of these three classes. The three groups had no significant differences in demographic characteristics, including age, sex, smoking status, BMI, waist circumference, and MetS and MetS ranking. In contrast, four ACEs, sexual abuse, and total ACEs were elevated in both MDD clusters compared with healthy controls. Significant differences between the three groups were observed for lifetime suicidal ideation (LT-SI), lifetime suicidal behaviors (LT-SB), total suicidal behaviors (Total-SB), and recurrence of illness (ROI). These scores increased from controls to cluster 1 to cluster 2. Significant differences were also observed across the three groups in current-SI, as well as OSOD and melancholia and physiosomatic scores. These differences remained significant after FDR correction and consistently showed increases from controls to cluster 1 to cluster 2. This pattern shows strong similarities with previous cluster analyses performed in Thai (19, 20) and Brazil (7, 16) patients with MDD and in analogy with these studies we labeled cluster 2 “MDMD” and cluster 1 “SDMD”. Thus, the discrimination of MDMD versus SDMD is externally validated by other clinical constructs such as ROI and lifetime and current SB scores.

In terms of immune-inflammatory markers, transferrin and albumin levels were significantly lower in MDMD and SDMD patients than in controls. M1 macrophage profile, Th17, Th2, IRS, CIRS, TNF signaling, and EGF were significantly higher in MDMD than in SDMD. Th17, CIRS, EGF and sCD40L were significantly higher in MDMD than in controls. TNF signaling was lower in SDMD than in controls.

Our observations indicate that 92 MDD patients received treatment with antidepressants, 10 were administered mood stabilizers, 44 were treated with atypical antipsychotics, and 68 were prescribed benzodiazepines. The administration of these drugs did not yield any significant effects on the immune variables, even in the absence of FDR p-value correction. Moreover, within the cohort experiencing MDD, no significant correlations were observed between the utilization of those medications, and current suicidal ideation, melancholic states, psychosomatic symptoms, and obsessive-compulsive disorder. There existed a significant correlation between the utilization of mood stabilizers and ROI, with a correlation coefficient of r = 0.227, p = 0.003. Nevertheless, the correlation in question ceased to be significant following the application of the FDR p-correction. Nonetheless, it is likely that this trend indicates that patients exhibiting elevated ROI are administered mood stabilizers.

### Correlations between ACEs, ROI, and current phenome

**Table 3** shows the correlations between total ACEs, the number of depressive episodes, ROI, and key symptom domains of the index episode. In the total sample (n = 165), ACEs were significantly positively correlated with ROI, Total-SB, Current-SI, melancholia, physiosomatic, and OSOD scores. Additionally, the number of depressive episodes and ROI showed strong correlations with the same features. In the restricted MDD subgroup, the results were consistent with those of the total sample, although the correlations were slightly weaker. **Figure 1** shows the partial regression of the OSOD score on the ROI score in the restricted MDD sample.

**Figure 1.**
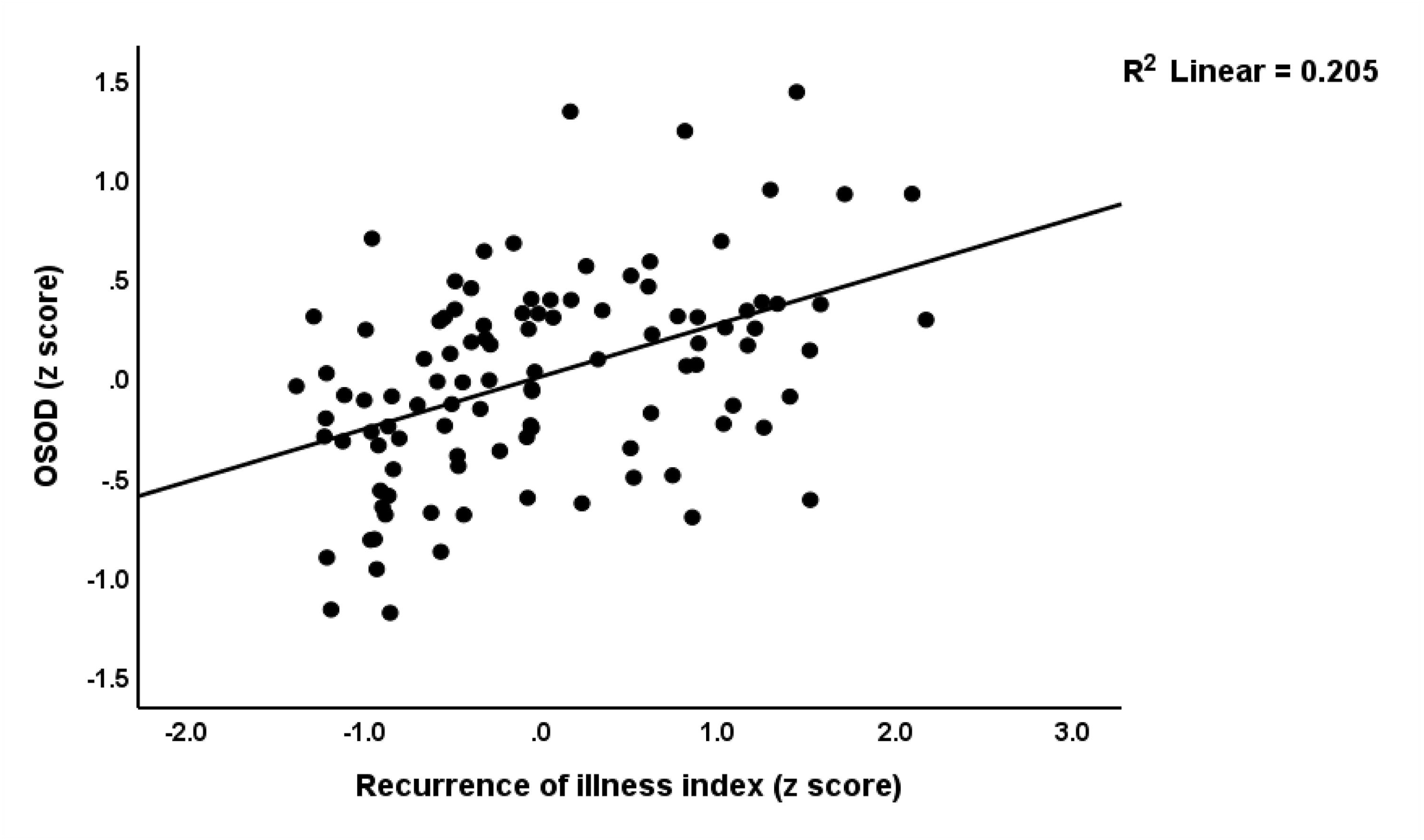
Partial regression of the overall severity of depression (OSOD) score on the recurrence of illness (ROI) score in the restricted MDD sample (p < 0.001).

### Multiple regression analysis with clinical scores as dependent variables

To examine the associations between different phenome scores and the combined effects of biomarkers and ACEs, we performed multiple regression analysis with clinical scores as dependent variables. **Table 4**, model #1 shows that total ACEs and Th17 profile (positively correlated), along with albumin and Flt-3 ligand (negatively correlated), explained 29.6% of the variance in the number of depressive episodes. Model #2, which included only MDD patients, shows that total ACEs and Th17 profile (positively correlated), Flt-3 ligand (negatively correlated), and female gender explained together 17.7% of the variance in the number of depressive episodes. Model #3 shows that total ACEs and Th17 profile (positively correlated), along with albumin (negatively correlated), explained 37.4% of the variance in ROI. In the restricted group of MDD patients, total ACEs and Th17 profile (positively correlated) explained 23.9% of the variance in ROI (see model #4).

**Table 4.**
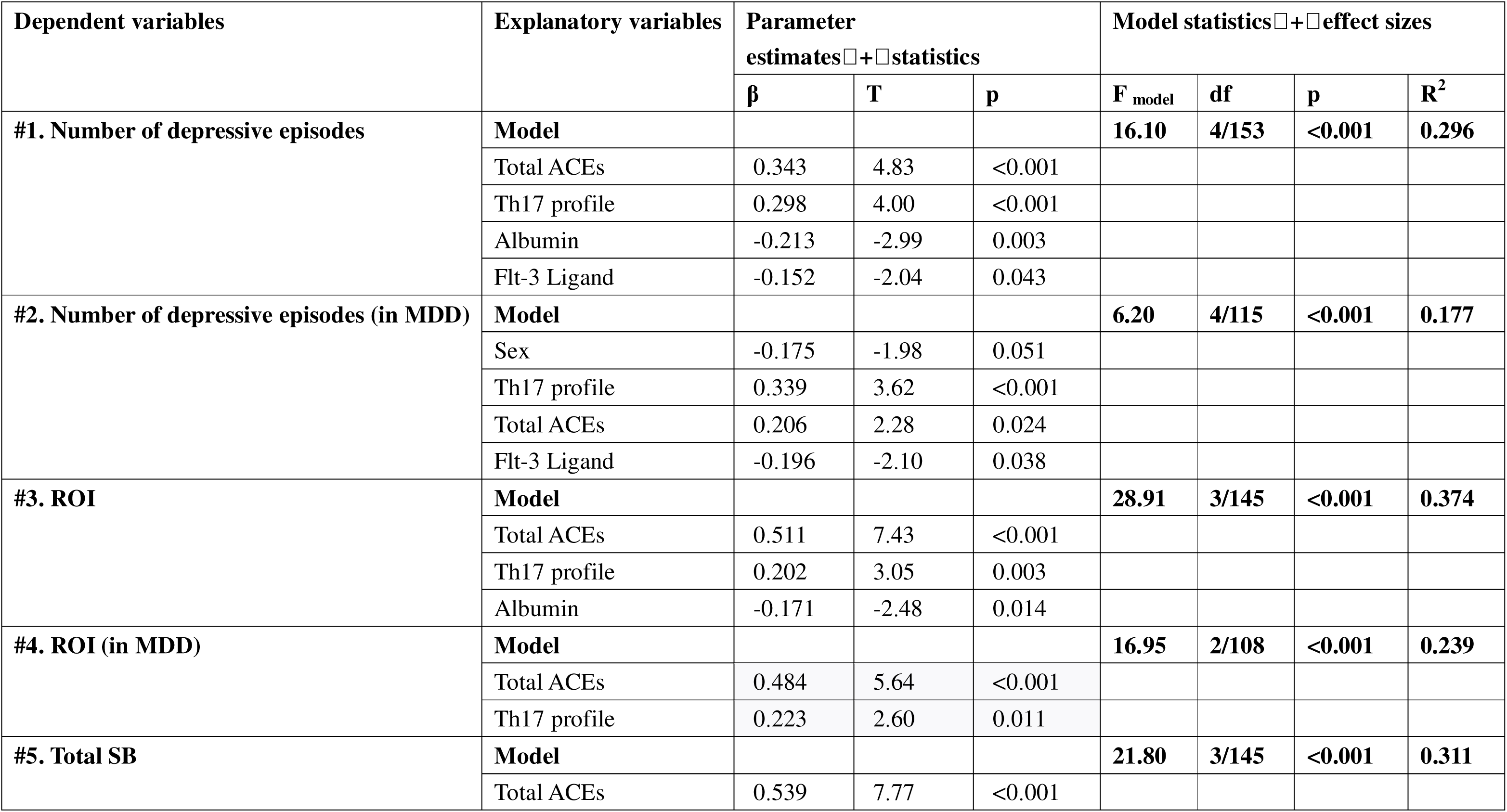
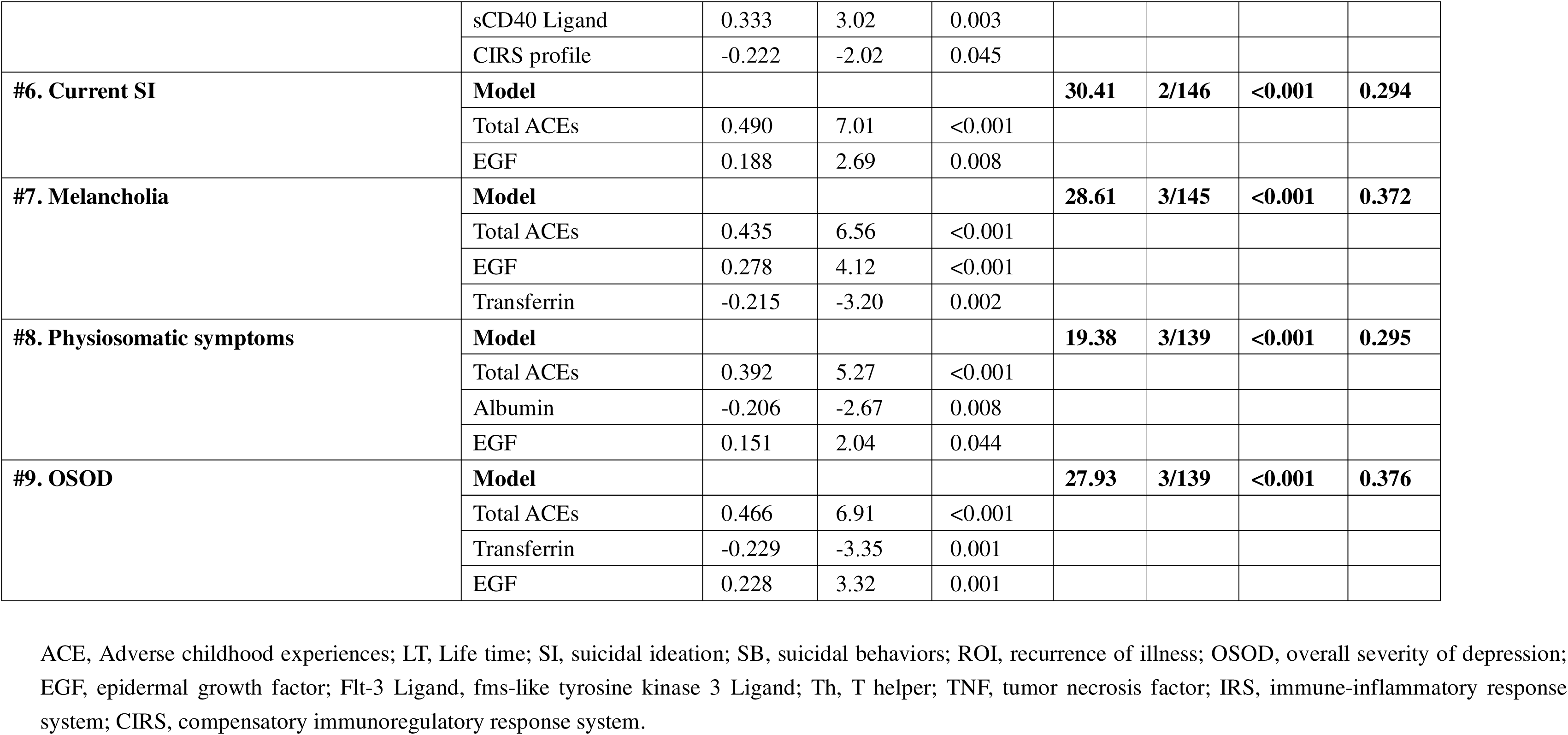
Results of multiple regression analyses with clinical data as dependent variables, and adverse childhood experiences (ACEs) and biomarkers as explanatory variables.

Model #5 shows that total ACEs and sCD40L (positively correlated), along with CIRS profile (negatively correlated), explained 31.1% of the variance in SB. Model #6 shows that total ACEs and EGF (positively correlated) explained 29.4% of the variance in current SI. Model #7 shows that ACEs and EGF (positively correlated), and transferrin (negatively correlated), explained 37.2% of the variance in the melancholia score. Model #8 shows that total ACEs and EGF (positively correlated), along with albumin (negatively correlated), explained 29.5% of the variance in the physiosomatic score. Finally, model #9 shows that total ACEs, EGF (positively correlated), and transferrin (negatively correlated) explained 37.6% of the variance in OSOD.

Partial regression (adjusted for age, sex, BMI) analysis showed that immune activation (constructed as a z composite score based on z EGF + z sCD40L + zTh17 + z CIRS – z albumin - z transferrin) was significantly associated with OSOD (r = 0.379, p < 0.001), melancholia (r = 0.397), and the physiosomatic score (r = 0.279, p = 0.003). This index was also significantly correlated with vegetative (r = 0.458, p < 0.001), current SI (r = 0.248, p < 0.001), affective symptoms (r = 0.301, p < 0.001), and ROI (r = 0.339, p<0.001). **Figure 2** shows the partial regression of OSOD on this immune activation index.

**Figure 2.**
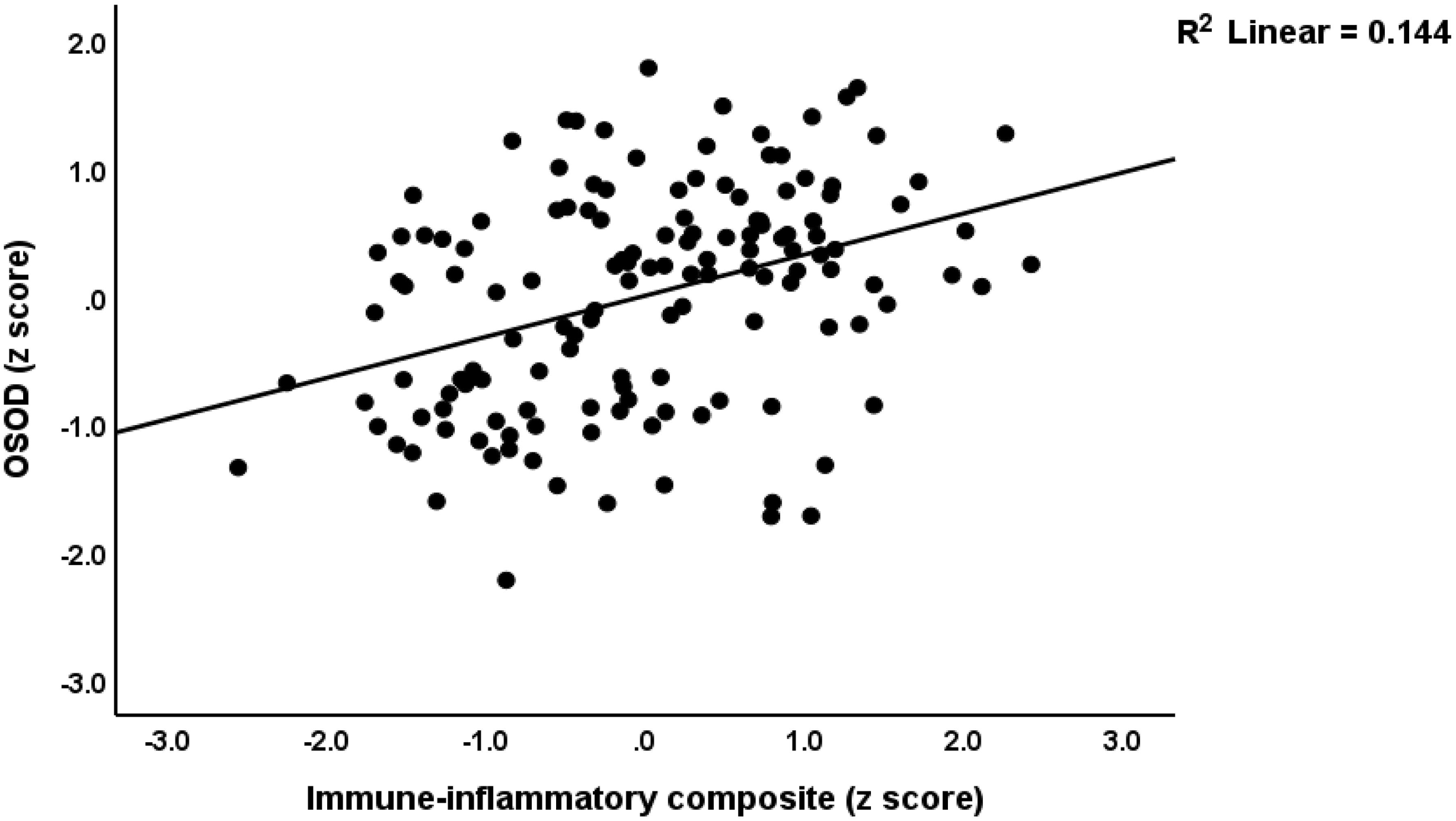
Partial regression of the overall severity of depression (OSOD) on a composite reflecting immune-inflammatory activation (constructed as a z composite score based on z EGF + z sCD40L + z Th17 + z CIRS – z albumin - z transferrin) (p < 0.001).

### External validation of the MDMD cluster by biomarkers

We have also examined the biomarker and ACE features of MDMD versus SDMD using binary logistic regression. The major aim of these analyses was to determine the accuracy of discriminating MDMD versus SDMD. **Table 5** presents the results of binary logistic regression analyses using MDMD as the dependent variable, with the SDMD group as the reference group. Analyses were conducted separately in all participants and in those without MetS to exclude putative effects of MetS.

**Table 5.** Results of binary logistic regression analysis with major dysmood disorder (MDMD) or major depression (MDD) as dependent variables.

| Dichotomies |  | Explanatory Variables | B | SE | Wald (df=1) | P | OR | 95% CI |
| --- | --- | --- | --- | --- | --- | --- | --- | --- |
| **MDMD (reference group: SDMD)**<br>**In subjects with and without MetS** | **Model #1** | Flt-3 Ligand | -0.427 | 0.204 | 4.39 | 0.036 | 0.652 | 0.438; 0.973 |
|  |  | TNF signaling | 0.876 | 0.247 | 12.59 | <0.001 | 2.402 | 1.480; 3.897 |
|  |  | Th2 profile | 0.595 | 0.214 | 7.70 | 0.006 | 1.813 | 1.191; 2.759 |
|  |  | Four ACEs | 0.507 | 0.195 | 0.25 | 0.009 | 1.660 | 1.134; 2.430 |
| **MDMD (reference group: SDSMD)**<br>**in subjects without MetS** | **Model #2** | Flt-3 Ligand | -0.543 | 0.221 | 6.04 | 0.014 | 0.581 | 0.377; 0.896 |
|  |  | TNF signaling | 0.821 | 0.260 | 10.00 | 0.002 | 2.272 | 1.365; 3.783 |
|  |  | Th2 profile | 0.764 | 0.249 | 9.42 | 0.002 | 2.147 | 1.318; 3.497 |
|  |  | Transferrin | -0.393 | 0.195 | 4.06 | 0.044 | 0.675 | 0.461; 0.989 |
|  |  | Waist circumference | -0.458 | 0.227 | 4.06 | 0.044 | 0.633 | 0.405; 0.988 |
| **MDD (reference group: HC)**<br>**in subjects with and without MetS** | **Model #3** | EGF | 0.933 | 0.212 | 19.43 | <0.001 | 2.541 | 1.679; 3.847 |
|  |  | Th2 profile | -0.482 | 0.196 | 6.05 | 0.014 | 0.618 | 0.421; 0.907 |
|  |  | Transferrin | -0.559 | 0.174 | 10.38 | 0.001 | 0.572 | 0.407; 0.803 |
|  |  | Albumin | -0.878 | 0.207 | 18.00 | <0.001 | 0.416 | 0.277; 0.624 |
| **MDD (reference group: HC) in subjects**<br>**without MetS** | **Model #4** | EGF | 0.703 | 0.192 | 13.37 | <0.001 | 2.020 | 1.386; 2.945 |
|  |  | Transferrin | -0.606 | 0.192 | 10.01 | 0.002 | 0.545 | 0.375; 0.794 |
|  |  | Albumin | -0.937 | 0.237 | 15.67 | <0.001 | 0.392 | 0.246; 0.623 |

MDMD, major dysmood disorder; SDMD, simple dysmood disorder; HC, healthy controls; ACE: adverse childhood experiences; Mets: metabolic syndrome; EGF, epidermal growth factor; Flt-3 Ligand, fms-like tyrosine kinase 3 Ligand; Th, T helper; TNF, tumor necrosis factor.

Model #1 indicates that, among all participants, the TNF signaling pathway, Th2 profile, four ACEs (positive correlations), and Flt-3 ligand (negative correlation) are significant predictors of MDMD relative to SDMD (χ² = 36.040, df = 4, p < 0.001; Nagelkerke R² = 0.262). The model’s accuracy is 70.3% (sensitivity = 58.7%, specificity = 80.0%), with an AUC of 0.762 (see **ESF, Table 4**, for other quality model metrics). Model #2, for participants without MetS, shows that MDMD is significantly associated with the TNF signaling pathway, Th2 profile, transferrin (positive correlations), Flt-3 ligand, and waist circumference (negative correlations) as compared with SDMD (χ² = 39.755, df = 5, p < 0.001; Nagelkerke R² = 0.319). The model’s accuracy is 73.3% (sensitivity = 67.7%, specificity = 77.8%), with an AUC of 0.785. **Figure 3** shows the ROC curve discriminating MDMD from SDMD and that 67.7% of MDMD patients were correctly classified with a specificity of 81.5% (Youden’s index = 0.492).

**Figure 3.**
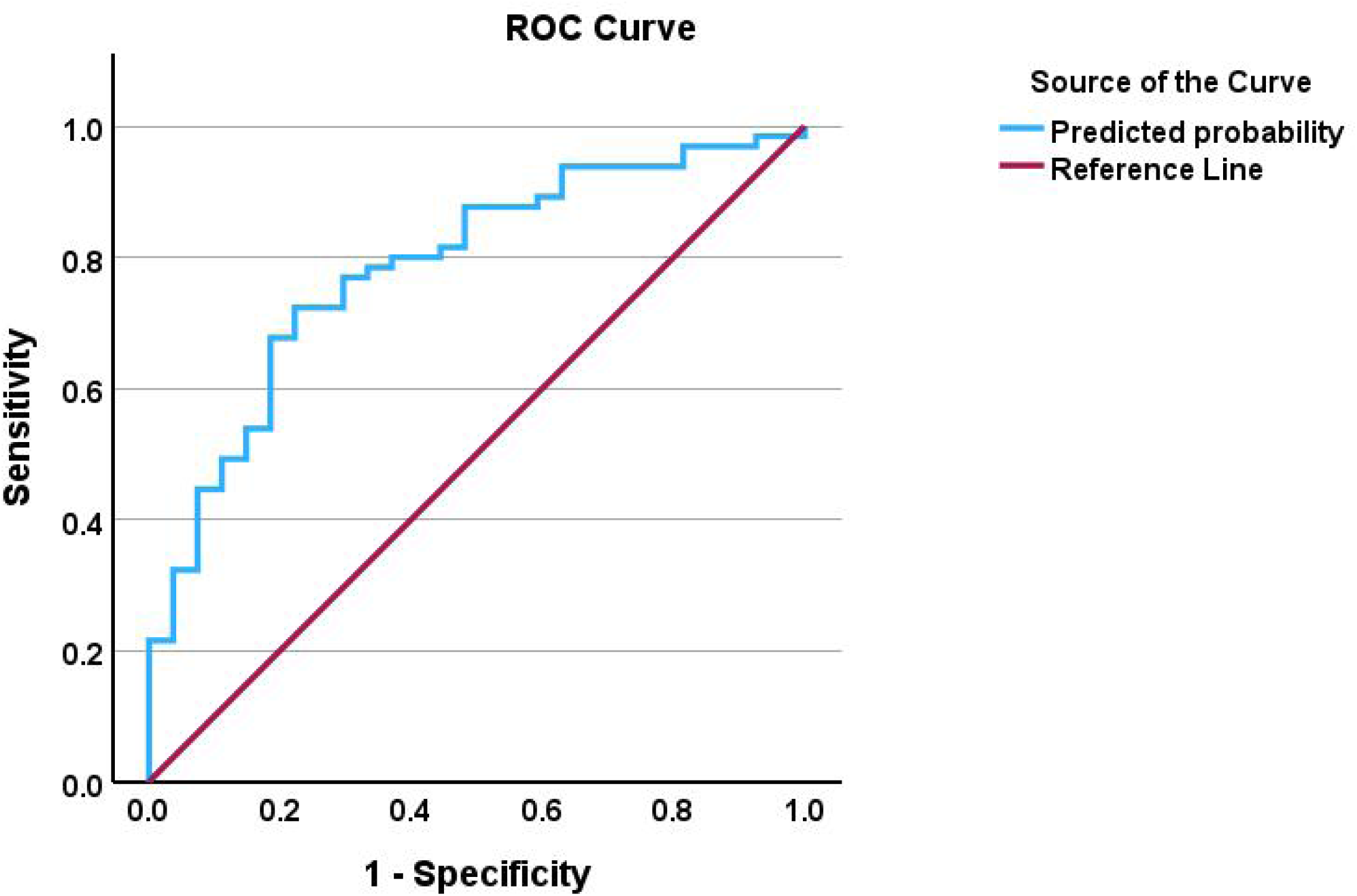
The receiving operating curve discriminating major dysmood disorder (MDMD) from simple dysmood disorder (SDMD).

### External validation of the MDD clinical diagnosis by biomarkers

We also examined the discrimination of MDD versus healthy controls using simple statistical analysis and multivariable binary logistic regression as well. **ESF, Table 5** presents the sociodemographic data of MDD patients and healthy controls. There were no significant differences in age, gender, education year, marital status, living environment, smoking, or drinking, and metabolic variables among the groups.

**Table 5** presents the results of binary logistic regression analyses using MDD as the dependent variable, with controls as the reference group. Model #3 indicates that, among all participants, MDD is significantly associated with EGF (positive correlation), as well as Th2 profile, transferrin, and albumin (negative correlations) (χ² = 83.584, df = 4, p < 0.001; Nagelkerke R² = 0.545). The model’s accuracy is 76.5% (sensitivity = 74.2%, specificity = 78.9%), with an AUC of 0.825. **Figure 4** shows the ROC curve discriminating MDD from controls. Youden’s index (0.521) shows that, at the most accurate cut-off value, 60.0% of the MDD patients were correctly classified with a specificity of 92.1%. Model #4, for participants without MetS, shows that EGF (positive correlation), transferrin, and albumin (negative correlations) are significant predictors of MDD (χ² = 65.402, df = 3, p < 0.001; Nagelkerke R² = 0.398). The model’s accuracy is 78.4% (sensitivity = 75.2%, specificity = 82.1%), with an AUC of 0.819.

**Figure 4.**
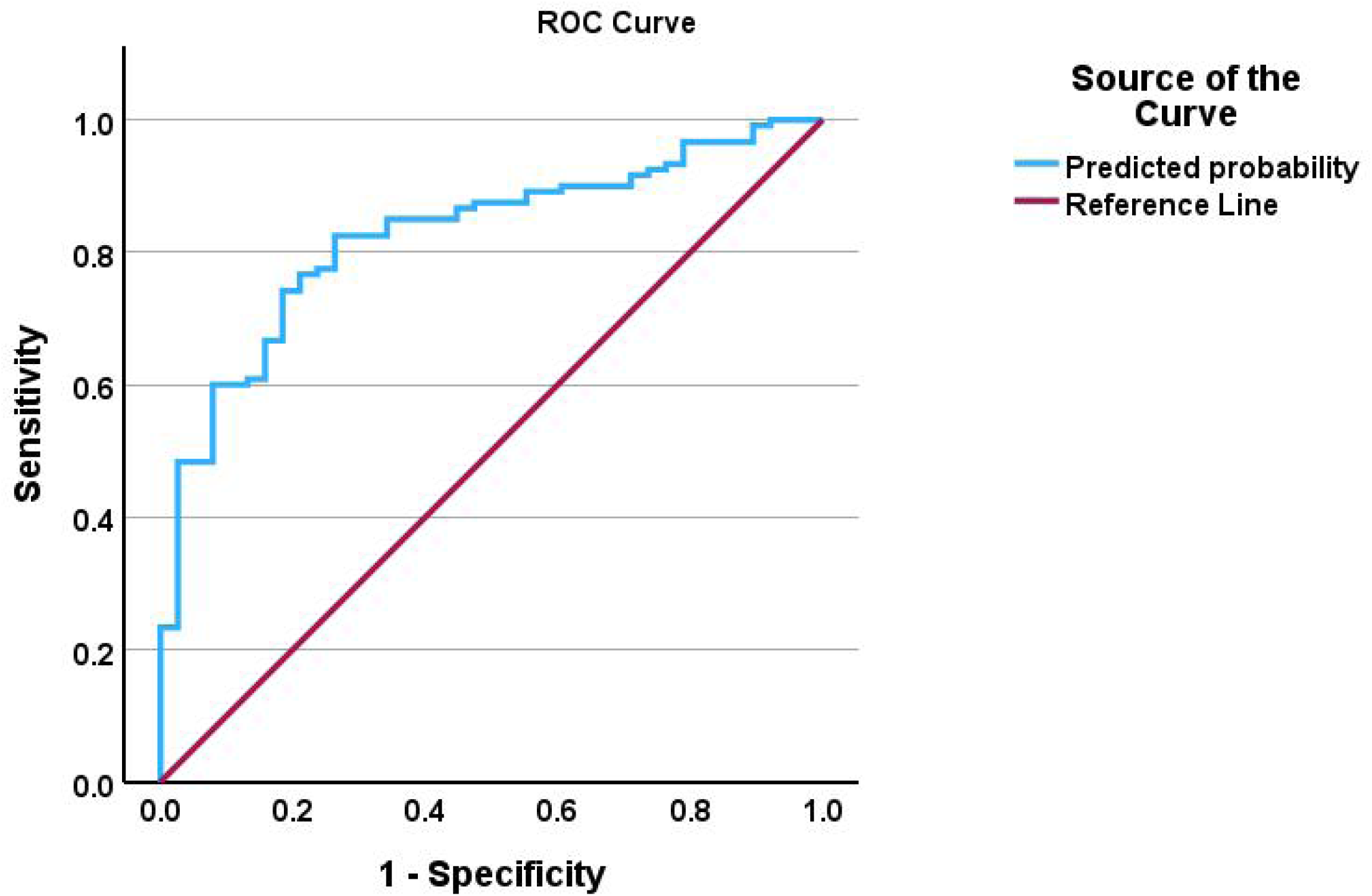
The receiving operating curve discriminating major depressive disorder (MDD) from healthy controls (HCs).

## Discussion

### Dissection of the phenome of depression in domains and subgroups

The first major finding of this study is that a bifactorial model most accurately represents the clinical manifestation of MDD, consisting of one general factor and one single group factor. All clinical phenome domains exhibited significant loadings on the general factor, suggesting that symptoms related to affection (depression, anxiety), physiosomatic symptoms, such as somatization, CFS-like symptoms, vegetative symptoms, and suicidal behaviors are intricately interconnected manifestations of a shared construct. This aligns with prior studies on those symptom dimensions in MDD (3–8). Previous research revealed a predominant general depression factor that extensively correlates with affective and somatic symptoms, with minimal residual specificity. Nevertheless, these results were based on bifactor analysis of a single rating scale consisting of affective and somatic symptoms. For example, a HAMD bifactor model revealed a strong general factor with extensive loadings on affective and vegetative items, alongside relatively weak specialized factors (49). The Center for Epidemiologic Studies Depression scale (CES-D) research similarly reveals a robust general distress component encompassing both affective and somatic elements, while affect was frequently maintained as a distinct factor (50). The Beck Depression Inventory (BDI)-II bifactor solutions demonstrated elevated general-factor loadings in mood, guilt, anhedonia, fatigue, sleep, and appetite, hence affirming minimal domain residuals (11, 51). PHQ-9 analyses utilizing bifactor-relevant indicators also identified a robust general factor encompassing cognitive-affective and somatic items (52). Overall, both self-report and observer scales endorse a general component that encompasses affective and physiosomatic symptomatology, with subdomains providing minimal additional variance.

Most importantly, we also identified a single group factor (SGF) which denotes the residual variance among physiosomatic symptoms—fatigue, CFS-like and autonomic symptoms, general physical complaints, and somatization—after accounting for OSOD. Consequently, it encapsulates variance specific to the physiosomatic symptoms that remain unaccounted for by OSOD. One bifactorial study found a general factor, and two subgroup factors, namely a somatic and a cognitive component (11). Other bifactor analyses of depression scales did not reveal a distinct, single group physiosomatic factor after the consideration of the general OSOD factor (10, 49–52). Previous models generally discovered several specifiers (e.g., cognitive–affective and somatic–affective) or observed that physiosomatic variance integrated within the overarching OSOD component. The discrepancies between these reports and our results can be explained by the wide range of physiosomatic symptoms assessed in the current study.

Depression universally has both affective and physiosomatic aspects, challenging the Western concept of mind and body dualism, as promoted by, for example, Rene Descartes in the 17^th^ century. Classical Chinese studies characterize depression as an integrated disruption of vital equilibrium, highlighting the interdependence of body and mind (53). Cross-cultural and primary care research indicates that chronic fatigue, pain, and somatic symptoms, are fundamental manifestations of emotional distress rather than distinct conditions (54). Contemporary cultural psychiatry advocates for a comprehensive biopsychosocial model, positing that affective and physiosomatic symptoms indicate the same fundamental dysregulation (55). This viewpoint was reconceptualized by Anderson et al. (56) who reported that depressed individuals suffer from concurrent co-expression of affective and physiosomatic dysregulation and that both dimensions show shared immune pathways (see also below). Consequently, these authors suggested that the term “psychosomatic” be replaced with “physiosomatic” in order to emphasize that these symptoms are rooted in an organic process (immune activation) (56). It is probable that physiosomatic symptoms are mediated by other pathways in addition to activated immune pathways, as they also rely on an orthogonal single group factor.

Based on our cluster analysis, we were able to classify MDD into two clinically distinct subgroups, namely MDMD and SDMD, with MDMD patients showing more severe symptoms of OSOD and physiosomatic symptoms, as well as suicidal behaviors. This novel classification of MDD into two subclasses is consistent with previous studies performed on Thai (19, 20) and Brazilian (7, 16) depressed samples. In fact, this classification into MDMD and SDMD is externally validated by increased ROI and suicidal behaviors in MDMD. This is consistent with previous research (21, 22). This distinction is further validated by immune biomarker findings (see Discussion, Chapter 3). Maes et al. (5) previously classified MDD into two qualitatively distinct categories: vital depression (which overlaps with melancholia) and non-vital depression. The significantly elevated melancholia score in MDMD compared to SDMD further substantiates this distinction. The melancholia MDD subtype is historically recognized as a more severe form, characterized by intense feelings of guilt, anhedonia, psychomotor disorders, circadian dysregulation, and vegetative symptoms (5, 57). Nonetheless, the present investigation revealed that MDMD/SDMD also demonstrates elevated ROI scores. Notably, research indicates that melancholic MDD exhibits a more severe trajectory, with a tendency for more rapid and frequent recurrences (57). All these findings suggest that the machine learning-generated class MDMD should be utilized by academics and clinicians as a distinct diagnostic class to signify a more severe subtype of MDD with a more severe trajectory.

### ACEs, ROI, and sensitization

In this study, we found a significant correlation between ROI and the depressive symptom phenotypes, OSOD, melancholia and physiosomatic symptoms. This indicates that as the number of depression episodes and lifetime suicidal behaviors increases, the severity of these factors rises in parallel with ROI. Previous studies conducted in Thailand and Brazil have confirmed that ACEs influence the severity of different symptom dimensions, with ROI potentially acting as a mediator of this effect (4–6, 14, 15). **Figure 5** depicts that when the number of episodes and ROI increases, there is a gradual increase in clinical severity of the symptom domains, indicating sensitization processes. Post’s hypothesis posits that with each depressive episode, the neurobiological thresholds for recurrence diminish steadily, a phenomenon referred to as kindling (58). Recurrent experiences sensitize limbic and stress-response circuits, enabling subsequent episodes to transpire independently, even in the absence of external stimuli.

**Figure 5.**
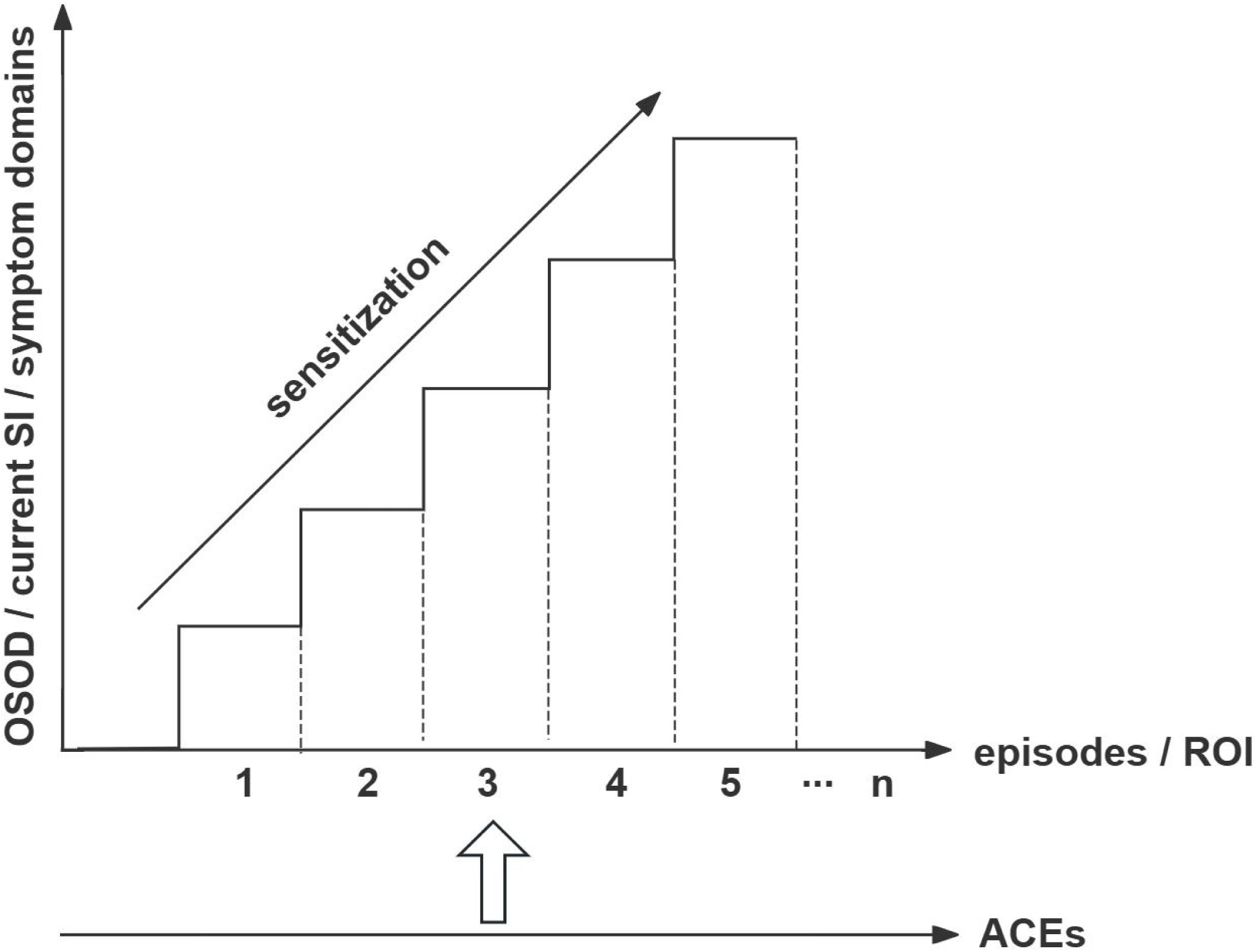
Correlations between adverse childhood experiences (ACEs), the number of depressive episodes, recurrence of illness (ROI), and different symptom domains of the index episode.

Several authors have proposed staging models for mood disorders, which classify the conditions into distinct stages based on various risk factors, including a family history of mental illness, patterns of early symptoms, self-reported recurrence of episodes, treatment resistance, and the level of functioning during non-episodic periods (59–61). The current DSM-5/ICD-10 diagnostic criteria for recurrent MDD are based solely on the binary classification differentiating between first-episode MDD from MDD with multiple episodes. These diagnostic approaches lack critical information about the lifetime trajectory of mood disorders, particularly the ROI, which is typically quantified using self-reported or physician-rated depression episode frequency, along with lifetime SI and suicide attempts (13). The absence of ROI also prevents the observation of sensitization markers in the disease processes, including NIMETOX biomarkers (1).

Therefore, the most appropriate method to classify and quantify depression is by using ROI, OSOD, the physiosomatic symptom domain, as well as MDMD and SDMD.

### Aberrations in immune profiles and clinical MDD features

Our results show that increasing ROI was associated not only with higher ACEs but also with increased Th17 signaling and lower albumin levels. In the restricted MDD sample, ROI was associated with Th17 signaling. Existing research has shown that with increasing ROI, the immune system in depressed patients becomes more sensitized (25). The phenomenon of time-dependent sensitization is well established, as exposure to stressors, whether psychological or organic in nature, progressively sensitizes the immune system (62–64). This immune sensitization is contingent upon re-exposure to stressors and gradually intensifies over time. Research by Maes et al. (20, 25, 65–70) demonstrated that with increasing ROI, this sensitivity is closely linked to the activation of various NIMETOX pathways including immune profiles, lipid metabolism disorders, and reduced antioxidant defenses and oxidative damage biomarkers.

At the same time, we found that immune system data can externally validate MDMD as compared with SDMD. Specifically, M1, Th17, IRS, Th2, CIRS, TNF signaling, and EGF were significantly higher in MDMD than in SDMD. In binary logistic regression analysis, we also found that TNF signaling, Th2 (positively correlated), and Flt-3L (negatively correlated) were significant predictors distinguishing MDMD from SDMD with great accuracy. Our previous studies in Thailand and Iraq have shown that MDMD is associated with higher expression of M1, Th1, Th2, Th17, IRS, CIRS, TNF signaling, and growth factors (19, 22). This reflects that the relationship between clinical phenotypic features and immune system characteristics is similar in China and other countries, suggesting that these changes are more universal.

Additionally, we found that these immune factors could be used to distinguish MDD from HC. EGF (positive correlation), Th2, transferrin, and albumin (negative correlations) significantly predicted MDD, with the model showing an accuracy of 76.5% (sensitivity = 74.2%, specificity = 78.9%), and an AUC of 0.825. This indicates that a large proportion of MDD patients exhibit immune system abnormalities: 60.0% of MDD patients were correctly classified with high specificity (92.1%). Compared to recent studies using serum high sensitivity C-reactive protein (hsCRP) levels >3 mg/L as the criterion for “inflammatory depression” or “immune-mediated depression,” approximately 25-30% of MDD patients showed inflammation (71). This suggests that measuring hsCRP alone is insufficient, and other immune factors should be considered to improve MDD diagnosis accuracy.

Most importantly, we discovered that, in addition to the imbalance between IRS and CIRS and the downregulation of negative APPs (transferrin and albumin), there are changes in isolated immune regulatory cytokines/growth factors (e.g., downregulation of Flt-3L and upregulation of EGF and sCD40L). Specifically, in multivariate regression analysis, we found that these depressive phenotypes could be jointly explained by the IRS activation and ACEs, with a high effect size. Th17, EGF, sCD40L (positively correlated), and CIRS, negative acute-phase proteins, and Flt-3L (negatively correlated) explained part of the variance. Suicidal behaviors were associated with CIRS, EGF and sCD40L.

Flt-3L enhances plasmacytoid DCs’ function, promotes Treg production, and supports the differentiation and maintenance of Treg and FoxP3+ Tregs through the secretion of tolerogenic cytokines (35, 36). Therefore, the decrease in Flt-3L suggests a reduction in Treg production, leading to a tolerance breakdown and lower immunoregulation. sCD40L activates T cells and immune responses by binding to CD40 on antigen-presenting cells (29). Thus, the increase in sCD40L in MDMD patients may lead to excessive immune activation. EGF regulates immune response by promoting epithelial cell proliferation, migration, and differentiation, which is crucial for tissue repair and homeostasis (33). However, excessive EGF expression can lead to immune system side effects, causing overactivation. EGF, by binding to the epidermal growth factor receptor, activates receptor tyrosine kinase activity and triggers multiple intracellular signaling pathways, including mitogen-activated protein kinase (MAPK), protein kinase B, and c-jun n-terminal kinase, promoting cell proliferation and gene expression (72). A similar condition was previously reported for other growth factors (VEGF, PDGF, FGF), which regulate immune responses through processes like cell division, endothelial cell chemotaxis, MAPK signaling, and angiogenesis (19). So, in MDMD, the increase in growth factors such as EGF might play a dual role in immune regulation, promoting repair while potentially triggering immune activation. In this regard, we observed that both increased and decreased CIRS (Th2) activities are associated with different depressive features. This is not contradictory; rather, it precisely reflects that patients with MDD are in a dynamic state of immune homeostasis imbalance (1, 24).

## Limitations

This study has a few limitations. Firstly, the cross-sectional design restricts the ability to make definitive conclusions about causality. Secondly, while we controlled for key confounders (e.g., metabolic variables), unmeasured variables (e.g., diet, physical activity, and genetic factors) may still have residual effects. For instance, we found that MetS, WC, and BMI did not seem to significantly impact the clinical phenome and immune system of MDD patients in this study. This contrasts with our findings in Brazil and a selected study sample reflecting the USA population with regard to obesity and MetS, where multiple interactions between increased MetS, BMI, and immune functions were observed in MDD (73, 74). Our cohort recruited in Chengdu, China, showed a MetS prevalence of 16.7% and an obesity rate of 11.0% (75), while in Western countries, such as the U.S., the MetS prevalence is as high as 36.9% (76) and the obesity rate reaches 40.3% (77).

## Conclusion

This study presents three key findings. First, through bifactor analysis, we successfully integrated multiple clinical domains in a general factor, introducing the concept of “OSOD” and extracting the single group factor of physiosomatic symptoms. Second, cluster analysis identified and confirmed two subgroups of MDD, namely MDMD and SDMD, which were cross-validated by increased ROI, suicidal behaviors and immune biomarkers, including increased TNF signaling and CIRS (Th2), a negative APP response, and lower Flt-3L. Third, we found that the ROI plays a key mediating role in modulating the relationship between ACEs and OSOD and physiosomatic symptoms, linked to enhanced immune activation (manifested by increased Th17 signaling and decreased albumin levels).

In conclusion, rather than relying solely on a singular bit of information (i.e., the binary diagnosis of MDD) and relying on a single rating scale score (for instance, the HAMD score), it is imperative for depression researchers to adopt a deep phenotyping approach. It is imperative that depression be articulated in accordance with the OSOD framework, as well as the physiosomatic domain, ROI, and the MDMD and SDMD constructs. **Figure 6** illustrates the relationships among these concepts and their relevance to immune disorders. Future research should continue to explore the complex relationships between immune system changes, symptom domains, and disease progression to lay the foundation for more personalized and effective treatment strategies for depression. Clinical psychiatrists should begin to employ these more significant concepts in order to gain a comprehensive understanding of the clinical picture.

**Figure 6.**
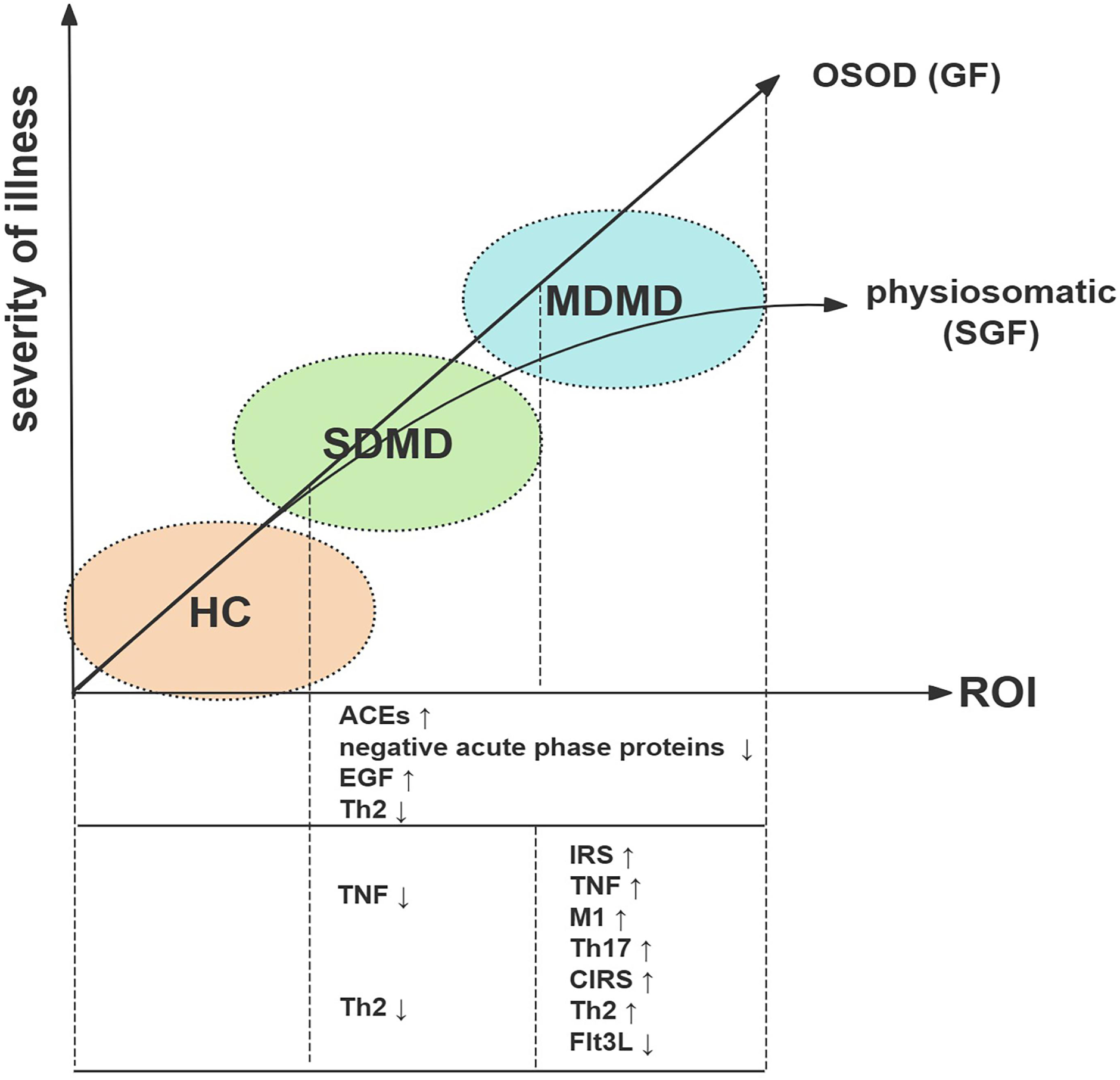
The associations between simple dysmood disorder (SDMD), major dysmood disorder (MDMD), overall severity of depression (OSOD) and the physiosomatic symptom domain. ROI, recurrence of illness; ACE, Adverse childhood experiences; EGF, epidermal growth factor; Th2, T helper 2; TNF, tumor necrosis factor; IRS, immune-inflammatory response system; M1, macrophage M1; Th17, T helper 17; CIRS, compensatory immunoregulatory response system; Flt-3 Ligand, fms-like tyrosine kinase 3 Ligand.

## Data Availability

The database created during this investigation will be provided by the corresponding author (MM) upon a reasonable request once the authors have thoroughly used the data set.

## Ethics approval

This study was approved by the ethics committee of Sichuan Provincial People’s Hospital approved the study [Ethics (Research) 2024-203] and strictly followed ethical and privacy regulations.

## Consent to participate

Before participating in this study, each subject provided written informed consent.

## Consent for publication

All authors have given their approval for this paper to be published.

## Declaration of Competing Interest

No conflict of interest was declared.

## Funding

This research was funded by the Health Science Research Project of Sichuan Province (Grant No.: ZH2024-203) and Sichuan Science and Technology Program “PIANJI” Project (Grant No.: 2025HJPJ0004).

## Author’s contributions

Mengqi Niu: visualization, writing - original draft, statistical analyses, recruiting participants. Yingqian Zhang: conceptualization, writing – review, editing, immune analyses. Xu Zhang, Chenkai Yangyang, Xiaoman Zhuang and Jing Li: recruiting participants. Michael Maes: supervision, conceptualization, formal analysis, writing - review and editing. All authors approved the submitted manuscript.

## Notes

### Competing Interest Statement

The authors have declared no competing interest.

### Author Declarations

This study was approved by the ethics committee of Sichuan Provincial People Hospital approved the study [Ethics (Research) 2024-203] and strictly followed ethical and privacy regulations.

