## Supplementary Table 1-5 for "Dissection of the clinical phenome of major depressive disorder into subdomains and subgroups in relation to activated immune-inflammatory profiles"

**ESF, Table 1.** Information on the construction of the clinical domain scores.

| **Items** | **Computation ways** |
| --- | --- |
| **Four ACEs** | PC extracted from emotional abuse, physical abuse, emotional neglect, and physical neglect. |
| **LT-SI** | PC extracted from six items of C-SSRS assessing suicidal ideation intensity and frequency. |
| **LT-SB** | Z Composite score. z (LT-SA) + z (LT-SI). LT-SA is the lifetime frequency of suicidal attempts. |
| **Current-SI** | A PC extracted from six items of C-SSRS assessing current suicidal ideation intensity and frequency. |
| **Total-SB** | Z composite score. z (LT-SB) + z (Current-SI). |
| **ROI** | Z composite score: z (number of depressive episodes) + z (frequency of suicidal ideation) + z (frequency of suicidal attempts). |
| **Pure HAMD** | Sum of depression, guilt, suicidal ideation, and loss of interest (Hamilton Depression Scale). |
| **Pure HAMA** | Sum of anxiety, tension, fear, and anxious behavior during the interview (Hamilton Anxiety Scale). |
| **Pure BDI** | Sum of Q1-Q10, Q12-Q15, and Q20 (Beck Depression Inventory-II). |
| **Pure chronic fatigue syndrome** | Z composite score: sum of the z scores of cognition (HAMA item) + muscle pain + muscle tension + fatigue + attention disorders + memory disorders + flu-like symptoms (the FibroFatigue Scale) + pain in the arms, legs or joints + feeling tired or lacking energy (SSS8) + concentration (BDI). |
| **Pure physiosomatic** | Z composite score: sum of the z scores of Q11 + Q12 + Q13 + Q14 + Q15 (HAMD) + Q7 + Q8 + Q9 + Q10 + Q11 + Q12 + Q13 (HAMA) + Q1 + Q2 + Q3 + Q9 + Q10 + Q11 + Q12 (FibroFatigue Scale) + Q1 + Q2 + Q3 + Q4 + Q5 + Q6 +Q7 (SSS-8). |
| **Vegetative symptoms** | Sum of anorexia, early awakening, mental retardation, loss of weight, and diurnal variation (Hamilton Depression Scale). |
| **Affective symptoms** | A PC extracted from pure HAMD, pure HAMA, STAI-state, and pure BDI. |
| **Melancholia score** | Z composite score: sum of z scores from HAMD (Q6 + Q8 + Q16 + Q17 + Q18) and BDI (Q4 + Q5 + Q6 + Q7 + Q8 + Q10 + Q12 + Q13 + Q18 + Q20). |
| Physiosomatic symptoms | A PC extracted from three somatic symptom domains which shape the single group factor in Table 1, namely SSS-8, pure CFS and pure physiosomatic |
| **OSOD** | Overall severity of depression, constructed as z score of a principal component from vegetative, affective, and physiosomatic symptom domains. |

PC, principal component, ACE: adverse childhood experiences, LT: lifetime.

**ESF, Table 2**. Overview of the immune system protein measured in the current study.

| **Protein abbreviations** | **Gene Symbol** | **> OOR (%)** | **Protein name / alias** |
| --- | --- | --- | --- |
| **Tf** | **TF** | 0 | Transferrin |
| **Alb** | **ALB** | 0 | Albumin |
| **EGF** | **EGF** | 0 | Epidermal Growth Factor |
| **Flt3L** | **FLT3LG** | 0 | FMS-like Tyrosine Kinase 3 Ligand |
| **CD40L** | **CD40LG** | 0 | CD40 Ligand or Tumor Necrosis Factor Superfamily Member 5 (TNFSF5) or CD154 |
| **IL-1α** | **IL1A** | 0.6 | Interleukin-1α |
| **IL-1β** | **IL1B** | 1.2 | Interleukin-1β |
| **sIL-1RA** | **IL1RN** | 0 | Soluble interleukin-1 receptor antagonist |
| **IL-2** | **IL2** | 0 | Interleukin-2 |
| **IL-4** | **IL4** | 0 | Interleukin-4 |
| **IL-5** | **IL5** | 96.3 | Interleukin-5 |
| **IL-6** | **IL6** | 0 | Interleukin-6 |
| **IL-10** | **IL10** | 0.6 | Interleukin-10 |
| **IL-12p70** | **IL12A, IL12B** | 0 | Interleukin-12 p70 |
| **IL-17A** | **IL17A** | 24.4 | Interleukin-17A |
| **IFN-α2** | **IFNA2** | 1.8 | Interferon-α2 |
| **IFN-γ** | **IFNG** | 0 | Interferon-γ |
| **TNF-α** | **TNF** | 0 | Tumor necrosis factor-α |
| **TNF-β** | **LTA** | 0 | Tumor necrosis factor-β or lymphotoxin-alpha (LT-α) |
| **TRAIL** | **TNFSF10** | 12.2 | TNF-related apoptosis-inducing ligand (TRAIL) or tumor necrosis factor ligand superfamily member 10 (TNFSF10) |

Adapted from:

*Almulla AF, Abbas Abo Algon A, Tunvirachaisakul C, Al-Hakeim HK, Maes M. T helper-1 activation via interleukin-16 is a key phenomenon in the acute phase of severe, first-episode major depressive disorder and suicidal behaviors. J Adv Res. 2024 Oct;64:171-181. doi: 10.1016/j.jare.2023.11.012. Epub 2023 Nov 13. PMID: 37967811; PMCID: PMC11464466.*

*Maes M, Rachayon M, Jirakran K, Sodsai P, Klinchanhom S, Gałecki P, Sughondhabirom A, Basta-Kaim A. The Immune Profile of Major Dysmood Disorder: Proof of Concept and Mechanism Using the Precision Nomothetic Psychiatry Approach. Cells. 2022 Mar 31;11(7):1183. doi: 10.3390/cells11071183. PMID: 35406747; PMCID: PMC8997660.*

**ESF, Table 3.** Description of the immune profiles used in this study.

| **Immune Profile** | **Members** |
| --- | --- |
| **M1 macrophage** | IL-1α, IL-1β, sIL-1RA, TNF-α, IL-6 |
| **T helper (Th)-1** | IFN-α2, IFN-γ，IL-2, IL-12p70 |
| **Th-17** | IL-6, IL-17A |
| **Th-2** | IL-4, IL-5 |
| **IRS** | All cytokines of M1 macrophage (except siL-1RA), Th1, and Th17 profiles |
| **CIRS** | IL-4, IL-5, IL-10, sIL-1RA |
| **TNF signaling** | TNF-α, TNF-β, TRAIL |

IRS: immune-inflammatory response system; CIRS: compensatory immunoregulatory system

Adapted from:

*Almulla AF, Abbas Abo Algon A, Tunvirachaisakul C, Al-Hakeim HK, Maes M. T helper-1 activation via interleukin-16 is a key phenomenon in the acute phase of severe, first-episode major depressive disorder and suicidal behaviors. J Adv Res. 2024 Oct;64:171-181. doi: 10.1016/j.jare.2023.11.012. Epub 2023 Nov 13. PMID: 37967811; PMCID: PMC11464466.*

*Maes M, Rachayon M, Jirakran K, Sodsai P, Klinchanhom S, Gałecki P, Sughondhabirom A, Basta-Kaim A. The Immune Profile of Major Dysmood Disorder: Proof of Concept and Mechanism Using the Precision Nomothetic Psychiatry Approach. Cells. 2022 Mar 31;11(7):1183. doi: 10.3390/cells11071183. PMID: 35406747; PMCID: PMC8997660.*

**ESF Table 4.** Performance evaluation metrics for different models (see Table 5).

| **Model** | **AUC** | **SE** | **Gini index** | **Max K-S** | **Overall model quality** |
| --- | --- | --- | --- | --- | --- |
| Model #1 | 0.762 | 0.038 | 0.524 | 0.467 | 0.69 |
| Model #2 | 0.785 | 0.038 | 0.570 | 0.501 | 0.71 |
| Model #3 | 0.825 | 0.027 | 0.650 | 0.562 | 0.77 |
| Model #4 | 0.819 | 0.032 | 0.639 | 0.610 | 0.76 |

AUC, area under the curve; SE, standard error; Max K-S, maximum Kolmogorov-Smirnov statistic

**ESF, Table 5.** Demographic data in patients with major depressive disorder (MDD) and healthy controls (HC).

| **Variables** | **HC (n = 40 )** | **MDD (n = 125 )** | **F/χ^2^** | **df** | **p** |
| --- | --- | --- | --- | --- | --- |
| Age (years) | 37.08 (13.75) | 35.69 (12.09) | 0.37 | 1/163 | 0.542 |
| Gender (Female/Male) | 27/13 | 87/38 | 0.06 | 1 | 0.802 |
| Education (years) | 13.88 (4.34) | 13.54 (3.33) | 0.26 | 1/163 | 0.613 |
| Married (yes/no) | 17/23 | 69/56 | 1.96 | 1 | 0.162 |
| Living (urban/rural) | 36/4 | 114/11 | FET | - | 0.760 |
| Smoking (yes/no) | 3/37 | 24/101 | FET | - | 0.091 |
| Drink alcohol (yes/no) | 7/33 | 12/113 | 1.86 | 1 | 0.173 |
| BMI (kg/m^2^) | 23.52 (4.07) | 22.30 (3.36) | 3.58 | 1/163 | 0.060 |
| Waist circumference (cm) | 79.38 (11.73) | 78.27 (11.32) | 0.28 | 1/163 | 0.595 |
| Mets (yes/no) | 10/29 | 22/102 | 1.17 | 1 | 0.279 |
| Ranking metabolic syndrome | 1.59 (1.46) | 1.42 (1.26) | 0.50 | 1/161 | 0.479 |
| Psychiatric history in family (yes/no) | 0/40 | 17/108 | FET | - | 0.013 |

Results are shown as mean (S.D.); F, results of analysis of variance; χ^2^, analysis of contingency tables; FET, Fisher’s exact probability test; BMI, body mass index; Mets: metabolic syndrome.
